# A Systematic Review of Interventions for Persons Living With Dementia: The Geriatric ED Guidelines 2.0

**DOI:** 10.1101/2025.02.28.25323113

**Authors:** Sangil Lee, Michelle Suh, Luna Ragsdale, Justine Seidenfeld, James D. van Oppen, Lauren Lapointe-Shaw, Carolina Diniz Hooper, James Jaramillo, Annie B. Wescott, Kaiho Hirata, Maura Kennedy, Lauren Cameron Comasco, Christopher R. Carpenter, Teresita M Hogan, Shan W. Liu, Geriatric ED Guidelines dementia writing group

**Affiliations:** Department of Emergency Medicine, University of Iowa Carver College of Medicine, Iowa City, IA; Section of emergency medicine, University of Chicago, Chicago, IL; Emergency Department, Durham VA Healthcare System, Clinical Associate, Department of Emergency Medicine, Duke University Hospital; Center of Innovation to Accelerate Discovery and Practice Transformation, Durham VA Health Care System; Centre for Urgent and Emergency Care Research, University of Sheffield, S1 4DA, UK; Department of Medicine, University of Toronto, Toronto, Canada; Faculdade Ciências Médicas de Minas Gerais, Belo Horizonte, Brazil; Frank H. Netter MD School of Medicine, Quinnipiac University; Galter Health Sciences Library & Learning Center, Feinberg School of Medicine, Northwestern University; Department of Emergency Medicine, International University of Health and Welfare Narita, Chiba, Japan; Department of Emergency Medicine, Massachusetts General Hospital, Harvard Medical School; Department of Emergency Medicine, Corewell Health William Beaumont University Hospital, Oakland University William Beaumont School of Medicine, Royal Oak, MI; Department of Emergency Medicine, Mayo Clinic, Rochester, MN; Department of Medicine Section of Emergency Medicine, Section of Geriatrics & Palliative Care, University of Chicago, Chicago, IL

**Keywords:** Dementia, Alzheimer’s Disease and Related Dementia, Intervention, Prevention, Emergency Department, Systematic Review

## Abstract

**Background:** The increasing prevalence of dementia poses significant challenges for emergency department (ED) care, as persons living with dementia (PLWD) more frequently experience adverse outcomes such as delirium, prolonged stays, and higher mortality rates. Despite advancements in care strategies, a critical gap remains in understanding how ED interventions impact outcomes in this vulnerable population. This systematic review aims to identify evidence-based ED care interventions tailored to PLWD to improve outcomes.

**Methods:** A systematic review was conducted in Ovid MEDLINE, Cochrane Library (Wiley), Scopus (Elsevier), and ProQuest Dissertations & Theses Global through September 2024. The review protocol was registered on PROSPERO (CRD42024586555). Eligible studies included randomized controlled trials, observational studies, and quality improvement initiatives focused on ED interventions for PLWD. Data extraction and quality assessment were performed independently by two reviewers, with disagreements resolved through discussion. Outcomes included patient satisfaction, ED revisits, functional decline, and mortality.

**Results:** From 3,305 screened studies, six met the inclusion criteria. Interventions included nonpharmacologic therapies (e.g., music and light therapy), specialized geriatric ED units, and assessment tools, such as for pain. Tailored interventions including geriatric emergency units and community paramedic care transitions were effective in reducing 30-day ED revisits and hospitalizations. However, heterogeneity in study designs and outcomes precluded meta-analysis. Risk of bias ranged from low to moderate.

**Conclusion:** This review underscores the urgent need for standardized and evidence-based interventions in ED settings for PLWD. Approaches including multidisciplinary care models and non-pharmacologic therapies demonstrated potential for improving outcomes. Future research should prioritize consistent outcome measures, interdisciplinary collaboration, and person-centered care strategies to enhance the quality and equity of ED services for PLWD.

**Key Points:**

1. Tailored interventions such as geriatric ED units and community paramedic care transitions significantly reduce ED revisits and hospital admissions among persons living with dementia.
2. Non-pharmacologic therapies, including music and light interventions, show potential for improving patient outcomes, though results are heterogeneous and require further validation.
3. The review highlights the urgent need for standardized protocols and interdisciplinary approaches to enhance emergency care for this vulnerable population.

**Why does this paper matter?:** This paper addresses critical knowledge gaps concerning emergency care for persons living with dementia, offering evidence-based insights to improve outcomes and guide the development of standardized, person-centered interventions in ED settings.

## Introduction

The number of older Americans (>65 years) with dementia is projected to increase from 7 million to 13 million between 2021 and 2050,^1^ while the global burden will triple over the same timeframe.^2^ Emergency departments (EDs) are often the first point of contact for individuals with dementia during acute health crises.^3–5^

People living with dementia (PLWD) are at heightened risk for preventable adverse emergency care outcomes including delirium, prolonged hospital stays, and increased mortality.^6^ They are also more likely to revisit the ED within 30 days.^3,7^ However, ED clinicians report a lack of training and resources to compassionately and cost-effectively manage PLWD during times of emergency,^8,9^ while emergency medicine-specific dementia detection strategies, best practices, communication strategies, and care transition approaches remain under-researched.^10–14^

The complexities and heightened risk associated with an ED visit for PLWD raise the need for more tailored approaches in emergency care. Suggested strategies to improve the experience or outcomes of ED care for PLWD include involving dementia care specialists to coordinate their care, engaging and communicating with patient’s care partners early in the episode of emergency care, implementing non-pharmacological interventions to manage agitation, and modifying the clinical environment to reduce stress and confusion.^13,15,16^

There remains a critical gap in measuring and quantitatively synthesizing how specific ED care processes for PLWD are effective. This study aims to fill this gap by systematically synthesizing the evidence linking ED care processes for PLWD with health outcomes. By identifying and analyzing these care practices, we aim to provide evidence-based recommendations to enhance the quality and safety of ED care for dementia patients. This systematic review was conducted as a part of the Geriatric ED Guidelines 2.0 effort. ^17^

## Methods

### Study overview

This is a systematic review and meta-analysis of studies describing ED interventions for PLWD. The study protocol is being reviewed at the PROSPERO (CRD42024586555). This review followed PRISMA 2020 statement guidelines.^18^

### Eligibility criteria

The following criteria for population, intervention, control, type of studies, and outcomes are listed below (Table 1).

**Table 1.** PICO question.

|  |  |  |
| --- | --- | --- |
| **Population:** | 65 years or older persons living with dementia or cognitive impairment presenting to the ED. | Excluded:<br>Non-human,<br>Simulation,<br>Index encounter in: non-ED outpatient, hospital, intensive care unit |
| **Intervention:** | Any intervention in target population in the ED |  |
| **Comparator:** | Usual care |  |
| **Outcomes:** | Primary: Patient or caregiver satisfaction | Secondary:<br>Patient/caregiver burden,<br>ED revisits,<br>ED length of stay,<br>Fall,<br>Functional decline,<br>Delirium superimposed dementia,<br>Mortality,<br>Hospital admission rates,<br>Hospital length of stay,<br>Sedation for agitation in the ED |

### Population

Persons aged 65 or older, living with dementia or cognitive impairment, presenting to ED were the population of interest. We defined dementia as a neurodegenerative process characterized by problems with memory, judgement, orientation, and executive function,^19^ and cognitive impairment as a condition that affects a person’s ability to think, learn, and remember, judge, and make a decision.^20^ The age cutoff of 65 was used as it was the common definition in the United States, but we allowed the age cutoff to be 60 to capture studies conducted elsewhere.^21^ Mild cognitive impairment (MCI) is considered the preclinical stage of dementia in this systematic review.^22^ Since our focus is on the dementia population, we included studies with more than 70% of the sample having dementia. Dementia was identified based on the past history, proxy history, risk stratification or assessment tools. Exclusion criteria included non-human, simulation, non-ED outpatient, hospital, intensive care unit (ICU) as a source of the index encounter. An Emergency Medical Services study may be included if the enrollment is in the ED.

### Intervention and Comparator

Studies were included if they described any intervention (including pharmacologic treatments) in the target population performed by physicians, nursing staff and pharmacists, pharmacologic interventions performed by physicians, nursing staff and pharmacists; furthermore, this intervention had to be compared to those receiving usual care. Studies describing screening through various risk scores, without any further intervention thereafter, were excluded.

### Type of study

Randomized control trials (RCT), quasi-experimental, observational study, pre- and post-study, and quality improvement studies were eligible for inclusion. Exclusion criteria included case reports, case series (n up to 5), review articles, scoping/systematic reviews, and qualitative studies.

### Outcomes

Patient or caregiver satisfaction was the primary outcome. Secondary outcomes included patient/caregiver burden, ED revisits, ED length of stay, fall, functional decline, delirium superimposed dementia, mortality, hospital admission rates, hospital length of stay, or sedation for agitation in the ED.

### Information sources

We searched the Ovid MEDLINE, Cochrane Library (Wiley), Scopus (Elsevier), and ProQuest Dissertations & Theses Global databases from inception to September 20, 2024. No language or other restrictions were applied.

### Search strategy and article selection

The search strategy is detailed in Appendix 1. A medical librarian created the electronic search strategy which was peer reviewed by another librarian who verified the validity and reproducibility.

### Data collection process

Database search results were uploaded to Covidence by the librarian. All entries from the searches were first deduplicated in EndNote and again in Covidence. During title/abstract screening and full text screening, each entry was screened by at least two independent reviewers in Covidence; disagreements were resolved by a third independent reviewer or through discussion in the review team meeting. Data were extracted using a standardized data collection form. Each study was extracted by two independent reviewers; discrepancies were resolved by a third reviewer or through discussion in the review team meeting.

### Data items

Data were extracted for author, year, study design and number of sites, country, study sample characteristics, dementia assessment tool (index or reference test), age (average and SD or IQR), sample size, % male/female, dementia prevalence (%), stage of dementia (if available, descriptive), place of living, comorbidities, frailty scale, ED length of stay. Outcome data included days in hospital, admission rate, ED revisit (time interval up to reviewer), mortality, patient/caregiver satisfaction (if they are reported), patient/caregiver burden, fall (in the ED, hospital, and after getting home), functional decline (after ED visit), delirium superimposed dementia after ED visits, sedation (in the ED), and agitation in the ED.

### Risk of bias assessment

The NewCastle Ottawa tool (cohort study version) for observational studies and the Cochrane risk of bias tool for RCTs were used. The quality of each study was assessed by two independent raters. The rating was provided for selection, comparability, outcome domains and a total score was used to classify them as low (Scores 0-3), moderate (Scores 4-7), and high risk of bias (8-10) for observational studies.(Appendix 4) The Risk of Bias 2.0 (RoB 2) tool (Appendix 5) assessed bias in randomized controlled trials across five domains: the randomization process, deviations from intended interventions, missing outcome data, outcome measurement, and selection of reported results. Each domain included signaling questions that guide judgments on bias risk, categorized as low risk, some concerns, or high risk based on a structured algorithm. The overall risk of bias was determined by aggregating domain-specific judgments, with a study rated as high risk if at least one domain is high or if multiple domains raise concerns. Otherwise, an online scoring guide was used for Cochrane risk of bias tools for parallel arm RCT and cluster RCT.^23,24^ Disagreements were resolved through discussion between the two raters or in the review team meeting until consensus was reached.

### Effect measures

We reported Absolute/Relative risk (RR), odds ratio (OR), risk difference, hazard ratio, and number needed to treat (NNT) for binary outcomes. These effects were extracted from the publications except NNT calculated from RR or OR using the online tool.^25^

### Synthesis

Meta-analysis was planned but not conducted due to the heterogeneity of identified study designs. Instead, we conducted a qualitative review of the selected studies.

## Results

### Study selection

A total of 3,309 studies were identified in September 2024. Four duplicates were removed. Of 3,305 remaining studies undergoing title and abstract screening, 3,250 were excluded for ineligibility. Fifty-five studies underwent full text review of which 6 were eligible and included (Figure 1).

**Figure 1.**
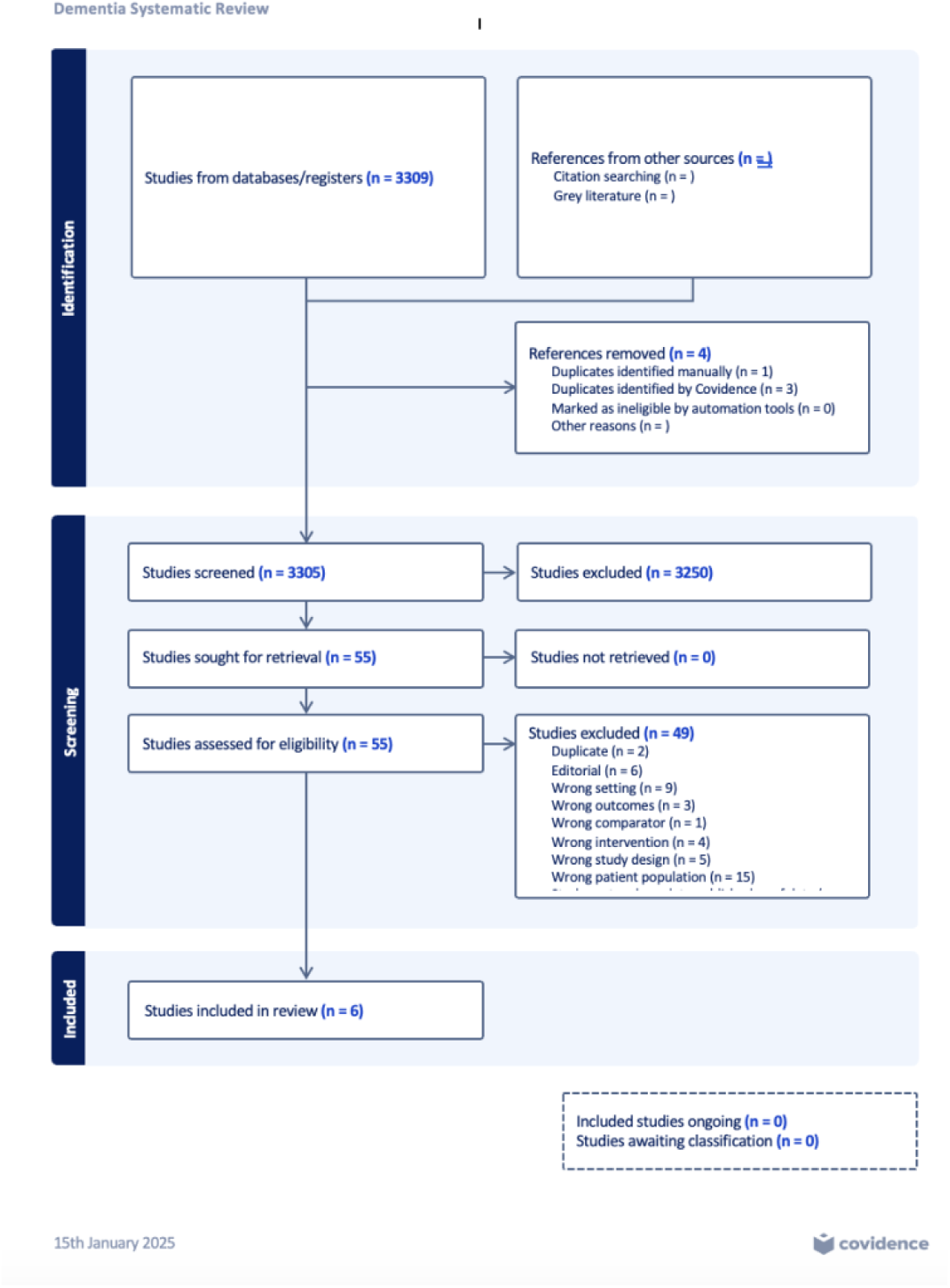
Selection of studies (PRISMA)

### Study characteristics

The characteristics of the 6 studies are listed in Table 2 and 3. Three studies were RCTs^26–28^ two were prospective observational studies^29,30^ and one was a historical cohort study.^31^ The study periods extended from 2018-2023. Two studies were conducted in the United States (Keene, Shah),^26,27^ two in Australia,^28,29^ and one study from France^31^ and one from Canada.^30^ Two studies determined dementia via the Six-Item Screener,^28,29^ two studies used medical record definitions,^31,32^ while Keene used the Short Blessed Test^33^ and Shah used the Blessed Orientation Memory Concentration Test.^27^

**Table 2.** Demographic summary of included studies.

| Author, Year | Design | Site | Country | Dementia assessment | Dementia prevalence | Sample size | Age | Sex, female% |
| --- | --- | --- | --- | --- | --- | --- | --- | --- |
| Keene, 2023<sup>26</sup> | RCT | 1 | USA | SBT | All SBT>4 | 151 | Median/IQR Music 84 (11), 2) Light 83 (8), 3) Music + light 83 (13), 4) Control: 84 (12) | Overall 61.7%, Music 51.5%, Light 60.6%, Music + light 65.7%, and Control 68.8% |
| Shah, 2022<sup>27</sup> | Subgroup of RCT | 3 | USA | BOMC | All BOMC>10 | 81 | Mean 78.4 ± SD 10.1 years. | Intervention 64%, Control 47% |
| Bosetti, 2020<sup>31</sup> | Non-RCT | 1 | France | DSM-V | 100% | 801 | Exposed group mean 87.0 (SD 5), Control group 86.0 (SD 6) | Exposure 73.5%, Non-exposure 71.6% |
| Fry, 2018<sup>28</sup> | RCT | 8 | Australia | SIS | 100% | 602 | Intervention group: Median 86 (IQR 79–90); Control group: Median 83 (IQR 74–89). | Intervention 70%, Control 72% |
| Beauchet, 2022<sup>30</sup> | Non-RCT | 1 | Canada | MCD from record review | 100% | 365 | Intervention mean 86.7 (SD 5.2), Control mean 86.6 (SD 5.4), | Exposure 69.8%, Non-exposure 63.1% |
| Fry, 2018<sup>29</sup> | Non-RCT | 4 | Australia | SIS | 77% | 181 | Mean 85.5 ± SD7.5 years | Overall 63.0% |

SBT: Short Blessed Test, BOMC: Blessed Orientation Memory Concentration Test, DSM: Diagnostic and Strategic Manual, SIS: Six item screening, MCD: Major cognitive disorder, SD: Standard deviation, IQR: Interquartile ratio. Stage of dementia, Place of living, and Comorbidities are not listed as more than half of the studies did not report them.

**Table 3.** Intervention and Outcome Table.

| Study Author, Year | Population | Intervention | Comparator | Outcome |
| --- | --- | --- | --- | --- |
| Keene, 2023<sup>26</sup> | Admitted older adults | Music, light, and both | Usual care | DSD: Music) 2/33 (RR 0.27, 95% CI 0.06-1.23, NNT6.26), Light) 3/33 (RR 0.41, 0.06-1.23, NNT 7.75), Both) 8/33 (RR 1.04, 0.42-2.55, NNT NA), None) 7/32 (Reference) |
| Shah, 2022<sup>27</sup> | Discharged older adults, substudy | Community paramedicine coaches# | Usual care | Intervention group had 75% decreased odds of an ED revisit within 30 days compared to those in the control group (OR 0.25, 95% CI 0.07 to 0.90), absolute numbers were 13/36 (36.1%) controls and 9/44 intervention (20.5%), NNT 6.39. |
| Bosetti, 2020<sup>31</sup> | Older adults presenting during office hours | Geriatric unit (MUPA unit)@ | ED care | Incidence of 30-day readmissions: The MUPA unit intervention had an OR of 0.68 (95% CI 0.47-0.98), p=0.03. In univariate analysis, p=0.04. In the exposed group, 63 (15.8%) were readmitted after discharge. In the control group, 89 (22.2%) were readmitted, NNT 15.71. |
| Fry, 2018<sup>29</sup> (Observational) | Older adults presenting to ED, mostly admitted. | PAINAD tool | NRS | Median time to analgesia: 90.0 minutes (IQR: 54.3–166.5), NRS: Median 5.5 (IQR 3.0–8.0), moderate pain, PAINAD: Median 1.0 (IQR 0.0–3.2), mild pain. Analgesia Administration: 75% received analgesia, with 53% receiving opioids. Pain Intensity and Analgesia Relationship: Weak correlation (Pearson's R = -0.019 and -0.006), NNT NA. |
| Beauchett, 2022<sup>30</sup> | Older adults with MNCD, on stretcher, assessed with ER2 | Multicomponent recommendations by ER2 tool* | Usual care | Intervention period had less incident hospital admissions: Unadjusted model - OR 0.61, 95% CI 0.40-0.93, p=0.022; Adjusted model - OR 0.59, 95% CI 0.38-0.91, p=0.018, NNT 8.2 |
| Fry, 2018<sup>28</sup> (Cluster RCT) | Older adults, suspected long bone fracture, SIS higher than 4 | PAINAD tool | NRS | Time to first analgesic dose (Intervention: median 83 min, IQR 48–158; Control: median 82 min, IQR 41–147; p = 0.42). Analgesia Within 60 Minutes of arrival: Intervention: 28%; Control: 32% (p = 0.19), NNT 25. Sensitivity Analysis for Cognitive Impairment Group: Intervention: median 90 min; Control: median 103 min (p = 0.62). |

Types of interventions were very heterogeneous, preventing meta-analysis (Table 2 and 3). These included light and music therapy,^26^ community paramedic Care Transition Intervention (CTI),^27^ a multidisciplinary team specializing in care of elderly ED patients,^31^ a multicomponent intervention^30^ while two examined the Pain Assessment in Advanced Dementia (PAINAD) tool.^28,29^

## Results of individual studies

Keene et al.^26^ conducted a pilot randomized controlled trial at an urban academic ED to assess the feasibility of using light and/or music therapy in preventing hospital-associated delirium among older adults 65 years and older in the ED. Exclusion criteria included Emergency Severity Index of 1, inability to consent, isolation precautions due to suspected SARS-CoV-2 infection, being legally deaf, intoxication, or presentation with a psychiatric chief complaint. Consented patients were screened for cognitive impairment with the Short Blessed Test (SBT);^33^ patients were enrolled in the trial if they tested positive for potential cognitive impairment (SBT score >4). Patients discharged from the ED were ultimately excluded from the study, but expected disposition was not considered an enrollment criterion. The study randomized 133 participants (median age similar across arms) into four parallel groups: music, light, both interventions, and control. The study was not adequately powered as it was a pilot study. Music was available through a bedside wireless speaker with either a classical or non-vocal jazz repeatable 2-hour long playlist. Classical was the default option if the patient was not able to choose. Light therapy was provided by a bedside full spectrum lightbox that mimicked natural light at 6,500 K color temperature and 5,000 lux brightness. The Confusion Assessment Method was performed at enrollment by a research assistant and then repeated by an inpatient bedside nurse upon patient arrival to the floor. Among the groups, delirium incidence was lowest in the music-only group (2/33, RR 0.27, 95% CI 0.06–1.23) and light-only group (3/33, RR 0.41, 95% CI 0.12–1.46). The combined music and light group (8/35, RR 1.04, 95% CI 0.42–2.55) showed no advantage over control (7/32). While statistical significance was not achieved, results suggest potential benefits of these non-pharmacologic interventions for delirium prevention. The study demonstrated feasibility of implementing in the ED and patient tolerance by measuring the completion of interventions and adherence, setting the stage for larger investigations.

Shah et al.^27^ conducted a pre-planned subgroup analysis of a RCT to evaluate the effectiveness of an adapted version of Coleman CTI,^34,35^ where no transition coaching was provided while the participant was in the ED, in reducing ED revisits (preplanned analysis) among cognitively impaired older adults. The study included 81 participants with cognitive impairment, identified by scoring > 10 on the Blessed Orientation Memory Concentration,^33^ (mean age 78, 57% female) from three university-affiliated hospitals. Participants receiving CTI had a home visit 24-72 hours after ED discharge by a trained community paramedic and follow-up phone calls post-ED discharge. During home visits, paramedics coached participants on CTI’s four self-management pillars: patient follow up, medication self-management, knowledge of red flag symptoms and use of a personal health record. The intervention significantly reduced the odds of 30-day ED revisits (OR 0.25, 95% CI 0.07–0.90) compared to usual care. The CTI did not significantly improve outpatient follow-up or self-management behaviors but demonstrated promise for enhancing ED-to-home care transitions in this vulnerable population.

Bosetti et al.^31^ conducted a historical cohort study to evaluate the effectiveness of a Geriatric Emergency Medicine Unit, known as the MUPA unit, for managing older patients with neurocognitive disorders (NCD) in the ED. Patients were included who had a diagnosis of NCD by DSM V and a medical record containing the terms: Alzheimer’s, vascular, mixed or frontotemporal dementia, dementia with Lewy bodies, or severe NCD. The MUPA unit is composed of an interdisciplinary team of physicians, nurses and social workers who perform geriatric assessments and individualized care plans for patients. The study included 801 patients aged ≥75 years (mean age 87, 72.5% female), comparing outcomes between those treated in the MUPA unit (n=400) and those receiving standard ED care (n=401). Patients treated in the MUPA unit experienced a 35% lower rate of 30-day readmissions (15.8% vs. 22.2%, *adjusted* OR 0.65, 95% CI 0.46–0.94) after adjusting for confounders (age, living at home, history of falls, the Charlson Comorbidity Index, and diagnosis of fall risk). Although hospitalization rates were higher in the MUPA group (57.8% vs. 47.1%), the intervention demonstrated the potential benefits of a multidisciplinary geriatric approach for improving outcomes of 30-day readmission (primary outcome) in this vulnerable population.

Fry et al.^28^ conducted a cluster randomized controlled trial to evaluate the impact of the Pain Assessment in Advanced Dementia (PAINAD) tool^36^ on the time to analgesia for cognitively impaired older adults (≳ 65) in the ED with suspected long bone fractures. PAIDAD is a tool developed to assess pain in patients with advanced dementia and particularly useful in patients with aphasia. The study included 323 patients at intervention sites using PAINAD and 279 at control sites using standard pain assessment methods. Patients at all sites were administered the Six Item Screener (SIS)^37^ prior to pain assessment by bedside nurses. Cognitive impairment was defined as a SIS score less than 4. At intervention sites, PAINAD was used on patients with a SIS score less than 4. Time to analgesia was defined as time from ED arrival to first dose of parenteral or oral analgesia. The median time to analgesia was 83 minutes at intervention sites and 82 minutes at control sites (p=0.42). A sensitivity analysis showed a non-significant reduction of 13 minutes in time to analgesia for patients at intervention sites (90 vs. 103 minutes, p=0.62). While PAINAD did not significantly reduce time to analgesia, it demonstrated potential clinical utility in improving pain recognition in this vulnerable population. The study highlights the challenges of timely pain management for older adults with cognitive impairment.

Beauchet et al.^30^ conducted a pre-post intervention study to assess the impact of the “Emergency Room Evaluation and Recommendations” (ER2) tool,^38^ which is a validated clinical tool to screen older patients in the ED to identify those at high risk of hospitalization and longer length of stay (LOS) in the ED than those who are not at high risk. In addition to its assessment part, ER2 has a tailor-made intervention guide based on responses to ER2, these recommendations were: 1) medication review by ED physician and pharmacist; 2) discharge planning team; 3) availability of walking aid, fall risk identification and encouraging mobility; 4) reorientation to time and place, assistance with basic needs, and toileting. The study included 356 participants aged ≥75 years with major neurocognitive disorder (MNCD) visiting the Jewish General Hospital ED. MNCD was defined by presence of diagnosis in a patient’s medical chart. During the intervention phase, ER2-tool screening and tailored recommendations for ED staff after triage were compared to usual care, and resulted in a 39% reduction in hospital admissions (OR 0.61, p ≤ 0.022, adjusted for baseline covariates). The authors did not evaluate the rate of return ED visits. However, the LOS in the ED increased significantly, by approximately 4.28 to 5.56 hours (p ≤ 0.008). The study highlights ER2’s potential to prevent hospitalizations through improved geriatric care while acknowledging the trade-off of extended ED stays.

Fry et al.^29^ (ref, note 2nd fry article) conducted a multi-center observational study in four EDs to evaluate the reliability and utility of the PAINAD scale for assessing pain and improving analgesia in older adults (age mean 85.5, range 63 to 100) with cognitive impairment (CI) identified by SIS score of less than 4 at the bedside. The study included 181 patients (mean age 85, 63% female) with suspected long bone fractures. The PAINAD scale showed good internal consistency (Cronbach’s α = 0.80) and moderate correlation with the Numeric Rating Scale (NRS) for pain (r = 0.39). Despite lower median pain scores on PAINAD compared to NRS (1.0 vs. 5.5), the tool was implemented as a routine pain assessment tool during the study period, suggesting feasibility in this cohort. The study concluded that PAINAD is a possible alternative to NRS for pain assessment in older adults with CI but highlighted the need for broader integration of pain assessment with caregiver input and comprehensive assessment strategies.

### Risk of bias in studies

The risk of bias assessment for the three observational studies with the NewCastle-Ottawa scale showed that one study was rated as low risk of bias,^31^ otherwise two other studies^29,30^ were rated as unclear risk of bias (Figure 2). The RCTs were rated as high risk of bias for one study and some concerns for two studies (Figure 3). Based upon the low quantity of direct evidence, the non-uniform approach to identifying PLWD across the included studies, the lack of a single dementia intervention, and the overall high risk of bias for the included RCTs and non-RCTs, the overall GRADE level of certainty for any ED intervention was very low.^39^

**Figure 2.**
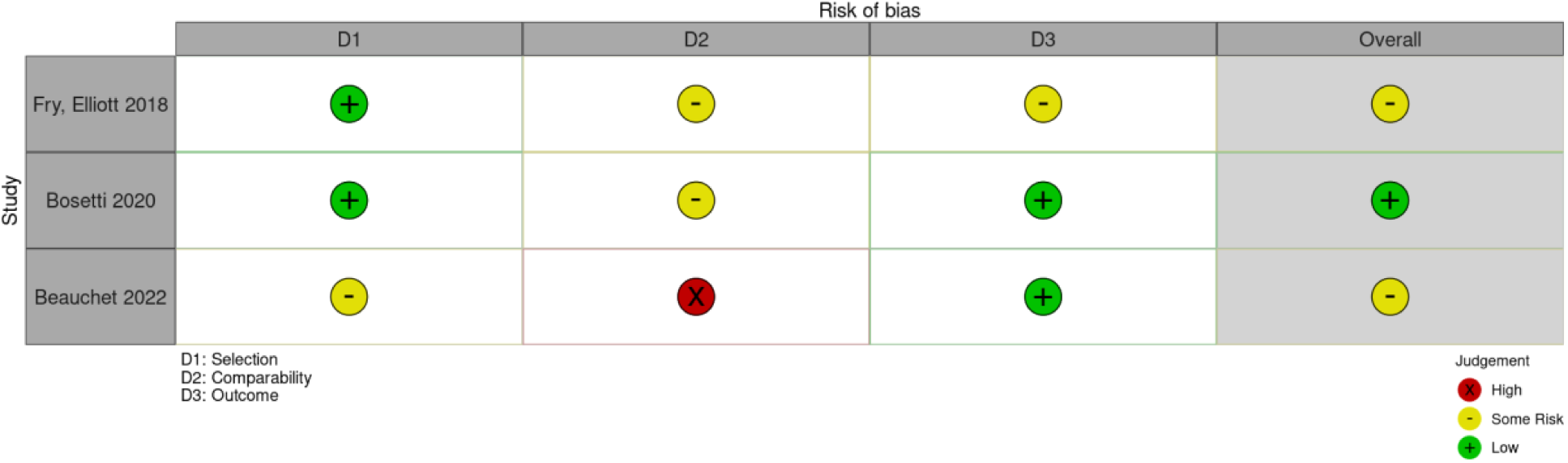
Risk of bias (cohort study) [Observational ROB](https://docs.google.com/spreadsheets/d/1v_kHZ68SB_Rv1q4AQZR82tK3VdGxik93OCBoX7oKH2I/edit?usp=sharing)

**Figure 3.**
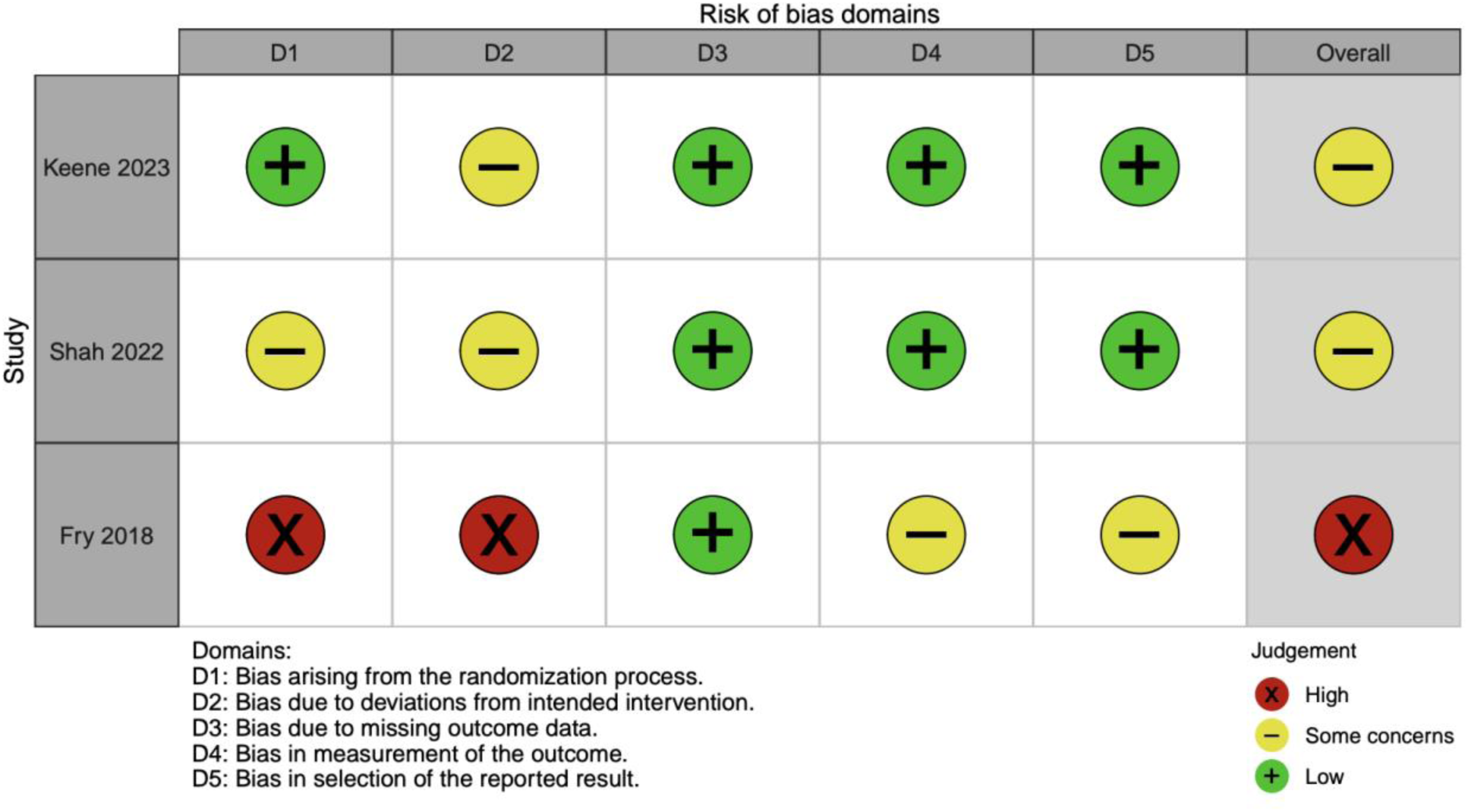
Risk of bias (RCT parallel and cluster) [RCT](https://docs.google.com/spreadsheets/d/1-qTo9SFdtXo8-uajcNyPeJct6raDzDXTu-Wpqx836d4/edit?usp=sharing)

## Discussion

Our systematic review identified a paucity of primary research evaluating interventions for PLWD in the ED, with six studies published thru September 2024.^26,28,29,31,32^ No two studies evaluated the same interventions which included: light and music therapy, community paramedic home visits, a multidisciplinary team specializing in care of older ED patients, a multicomponent intervention; and use of the Pain Assessment in Advanced Dementia (PAINAD) tool. None of these studies defined dementia in the same way, or evaluated the same dementia intervention. Interventions examined many outcomes other than those typically reported (e.g. ED readmissions and hospitalizations). Our systematic review highlights the need for more standard outcome measures, to include patient outcomes, process-oriented outcomes, and patient reported outcome measures.^40,41^

Despite the increase in incidence of dementia and ED visits by PLWD,^42^ the dearth of ED interventions designed to improve the experience, process, or outcomes of care is alarming. Although more than 10 years have passed since the initial Geriatric ED Guidelines^43^ were published and 20 years since the American Geriatrics Society Research Agenda Setting process prioritized “interventional trials should be designed to determine the effect on outcomes of better screening and management of cognitive impairment in older ED patients”,^44^ trial data is essentially non-existent. Patient stakeholders and multidisciplinary clinical groups identified various challenges associated with the ED care for PLWDs, including identifying and managing functional dependence, behavioral symptoms, and pain management using a unique tool or method to evaluate PLWDs.^13^ PLWDs may experience disparities in treatment and negative consequences, such as hospitalization rate,^45^ during ED visits. The heterogeneity of interventions evaluated and outcomes assessed, has previously prevented specific recommendations for ED interventions to improve outcomes for PLWD,^45^ and our systematic review has yielded similar conclusions. The Geriatric Emergency Care Applied Research Network 2.0 – Advancing Dementia Care (GEAR 2.0-ADC) is a research program aspiring to improve the experience, process, and outcomes of emergency care for PLWD.^10,12–14,16,46^ GEAR 2.0-ADC has identified research priorities for improving emergency care for PLWD and their care partners. ^10,12–14,16,46^ These priorities pertain to communication and decision making, dementia detection, ED care practices, and care transitions. Some recommendations for improving emergency care for PLWD include: evaluating for cognitive impairment,^47,48^ limiting the use of sedation and physical restraints,^49^ focusing on appropriate autonomy and maintaining functional capacity,^50^ adapting communication strategies and conscientious inclusion of care partners, building reliable infrastructure, and using transdisciplinary outcome measures.^51^ To address these issues, some experts suggest adopting a partnership approach between carers and ED nurses and developing research priorities through consensus-driven methods involving diverse stakeholders, including patients and care partners.^16,45^

Given the heterogeneity in interventions and outcomes for the included studies, we were unable to conduct a meta-analysis. However, two studies^30,31^ report interventions that decreased ED revisits^31^ and hospital admissions^30^ These interventions included the GEMU and ER2 disposition guide. Shah et al. also reported that a post-ED paramedic visit and a phone call follow-up decreased the rate of 7-day ED return, but not 14-day ED returns. (Shah et al. Cite) EDs are crucial sites of care for older adults, accounting for 18% of visits and 40% of admissions.^52^ However, the fast-paced ED environment often conflicts with the complex needs of older patients, particularly PLWD.^52^ Research priorities include developing geriatric-focused dementia-friendly EDs, improving pain assessment by using the appropriate tools for PLWD, and implementing care transition interventions. Non-pharmacological interventions, such as individualized music show promise in managing dementia-related behaviors.^53^ Overall, these papers highlight the need for tailored, evidence-based approaches to enhance ED care for older adults and dementia patients.^26–31^ The limited evidence from ED studies might be attributed to the number of studies from ED, funding stream, and regulation related to the institutional review board.^54,55^

The studies employed interventions that appeared reasonable and likely to be valued by both clinicians and patients. However, the anticipated benefits were not consistently observed when process measures such as increased hospitalizations and longer lengths of stay, were evaluated. This highlights the importance of adopting more standardized application of dementia assessment, reproducible interventions, and the selection of outcomes in the future. This systematic review evaluates the evidence for person-centered interventions in serious physical illness, finding mixed results on traditional outcomes but suggesting incorporating person-centered outcomes could provide a more comprehensive understanding.^56^ Incorporating person-centered outcomes alongside conventional service metrics could provide a more comprehensive view of impact of an intervention. Overall, we recommend increasing the consistency of ED intervention for PLWD for reproducibility and external validity, using rigorous study designs,^21^ consideration of underlying bias, and transparency around inclusiveness.Interventions should be evaluated with outcome measures that are relevant to patient outcome, such as mortality, 30-day readmission, and disposition to skilled nursing facility, and patient reported outcome measures. Future studies should focus on promising interventions, such as pain measurement, enhanced communications, reducing caregiver stress, the use of an ED activity cart, and reducing delirium superimposed on dementia.

### Limitations

First, it is possible that we did not identify an ongoing intervention study as we did not include grey literature or protocol registries in the strategy. Our search and selection process took place in 2024, and we intended to find all the studies that will have any results before this systematic review was concluded. Second, our review is limited in that literature based on qualitative research study was possibly excluded. Third, there may be interventions outside of the emergency care setting reported in the literature which were not considered but may be relevant for adaptation or implementation. Fourth, without a uniform approach to assessing presence or absence of dementia, each of our included studies may have included different proportions of dementia or stratifications of dementia severity. Lastly, even if all of the included studies used the same method of defining dementia’s presence or absence, relying on past history or proxy history is imperfect in ED settings and may have missed a large proportion of individuals with dementia.^57^

## Conclusion

Despite the growing incidence of dementia and the increasing burden on EDs, the evidence for effective, consistent, and reproducible interventions remains limited. This systematic review showed that few studies have examined the impact of ED-based interventions for PLWD on process outcomes and found no research exploring patient-oriented outcomes. The scant existing literature cannot support any confident management recommendations, thus highlighting the critical need for ongoing study of interventions to improve emergency care for PLWD. While some interventions demonstrated promising outcomes, such as reduced ED revisits and hospital admissions, significant variability in study design, interventions, and outcomes precluded a meta-analysis. This underscores the urgency for standardized protocols and robust research to address the unique challenges faced by PLWD in ED settings. Future efforts should focus on interdisciplinary collaboration, stakeholder engagement, and consensus-driven research priorities to establish evidence-based guidelines that enhance the safety, quality, and equity of emergency care for this vulnerable population. Our future steps include rating the level of certainty using the GRADE approach.

## Data Availability

All data produced in the present work are contained in the manuscript.

## Appendix 1. Search strategy

Appendix 1. Search strategy.
| Database searched | Date searched | Results |
| --- | --- | --- |
| Ovid MEDLINE(R) ALL <1946 to September 19, 2024> | 09/20/2024 | **2736** |
| Cochrane Database of Systematic Reviews Issue 9 of 12, September 2024 | 09/20/2024 | **5** |
| Cochrane Central Register of Controlled Trials Issue 8 of 12, August 2024 | 09/20/2024 | **541** |
| Scopus (Elsevier) | 09/20/2024 | **249** |
| ProQuest Dissertations & Theses Global | 09/20/2024 | **163** |
| Total |  | **3694** |
| After de-duplication |  | **3306** |

### Example Search Methods

The review authors partnered with a research librarian (ABW) to create a comprehensive search of the literature. The search was adapted from a previous search created as part of the original GEAR 2.0 scoping review projects. The search strategy combined database-specific controlled vocabulary and keyword searching related to the concepts of dementia in the emergency department setting. The search was conducted on September 20, 2024 in the following databases: Ovid MEDLINE, Cochrane Library (Wiley), Scopus (Elsevier), and ProQuest Dissertations & Theses Global. All databases were searched from inception to present without the use of filters or limits. Records were downloaded and underwent multi-pass deduplication in a citation management software (EndNote) and unique records were uploaded to Covidence for initial screening by two independent reviewers.

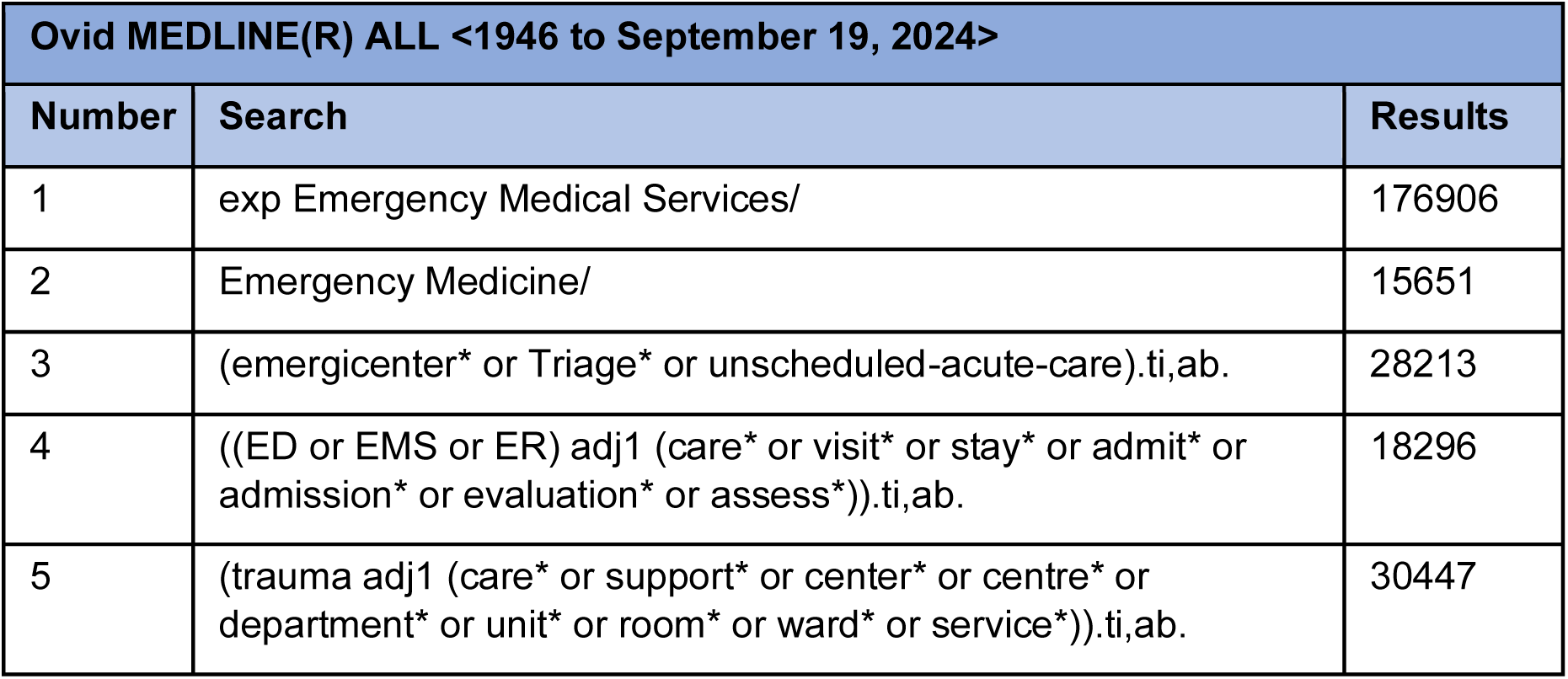
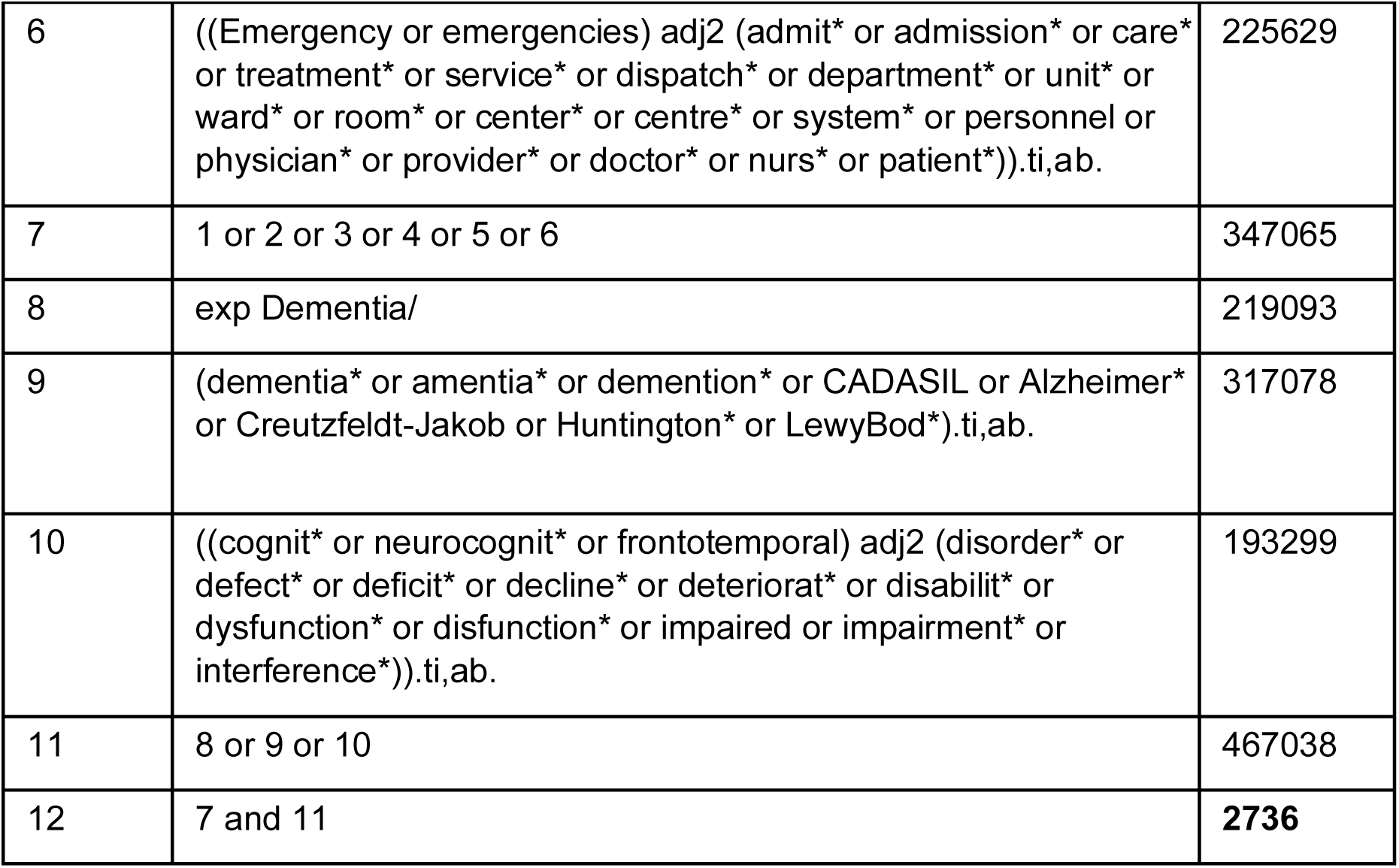
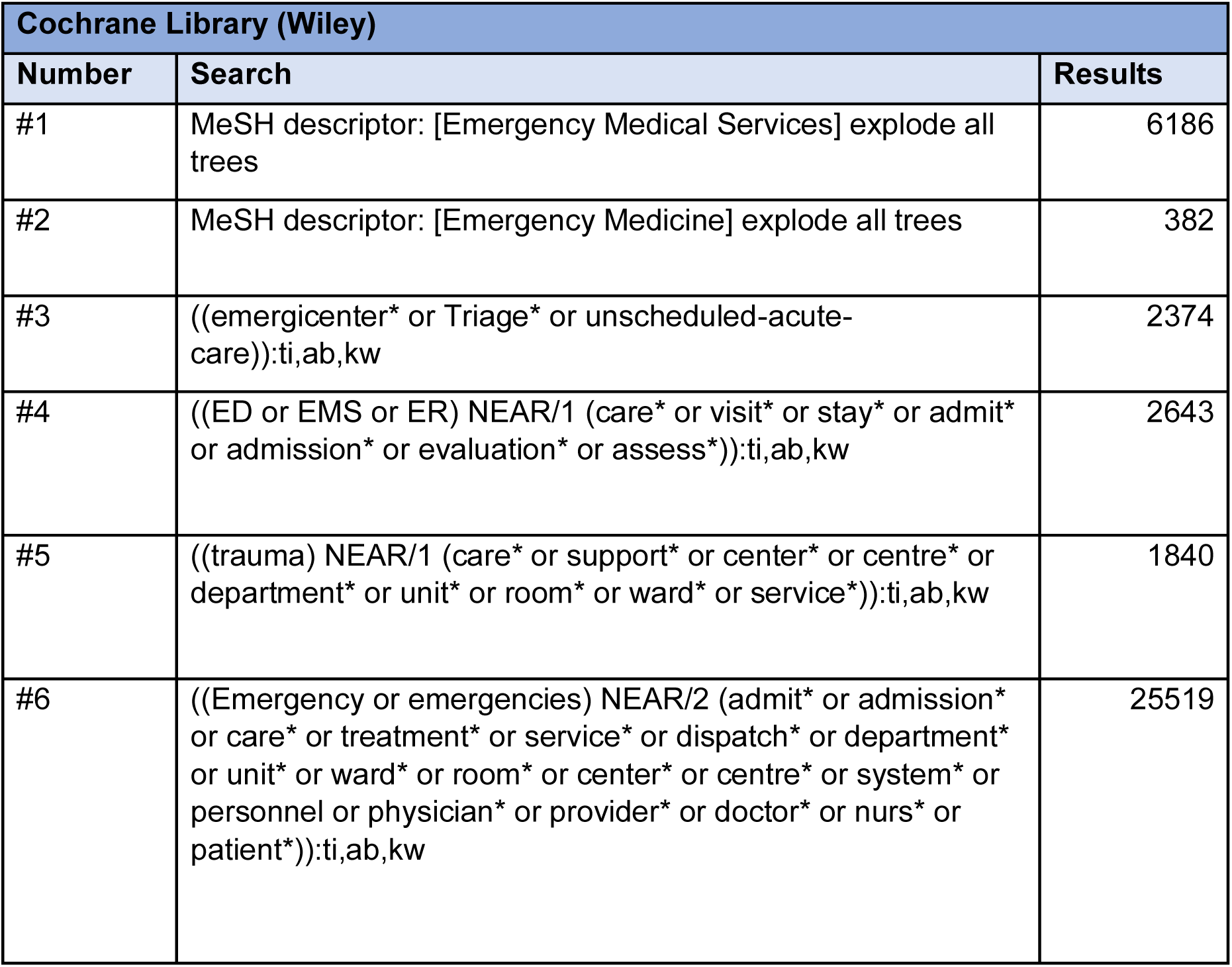
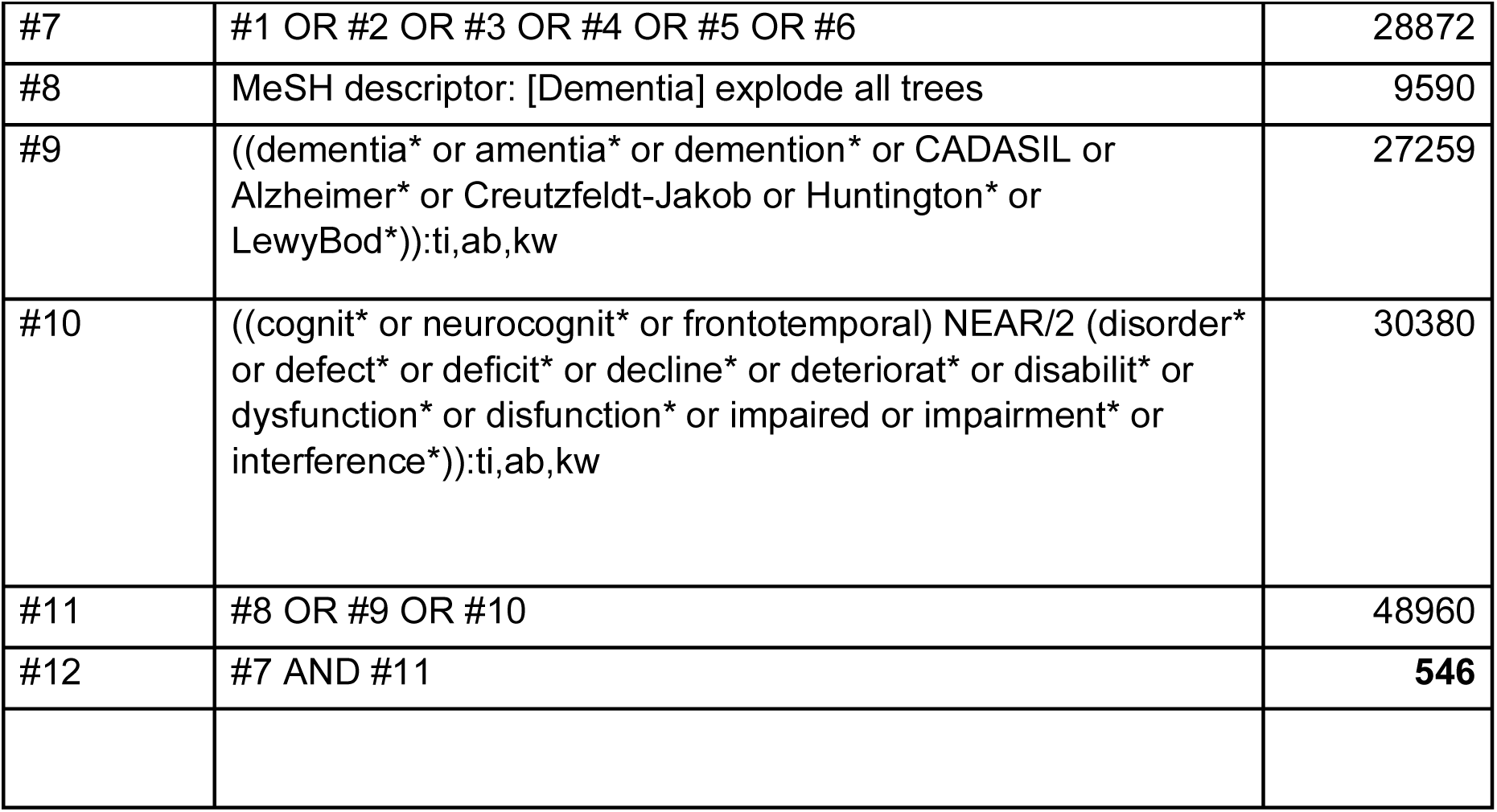
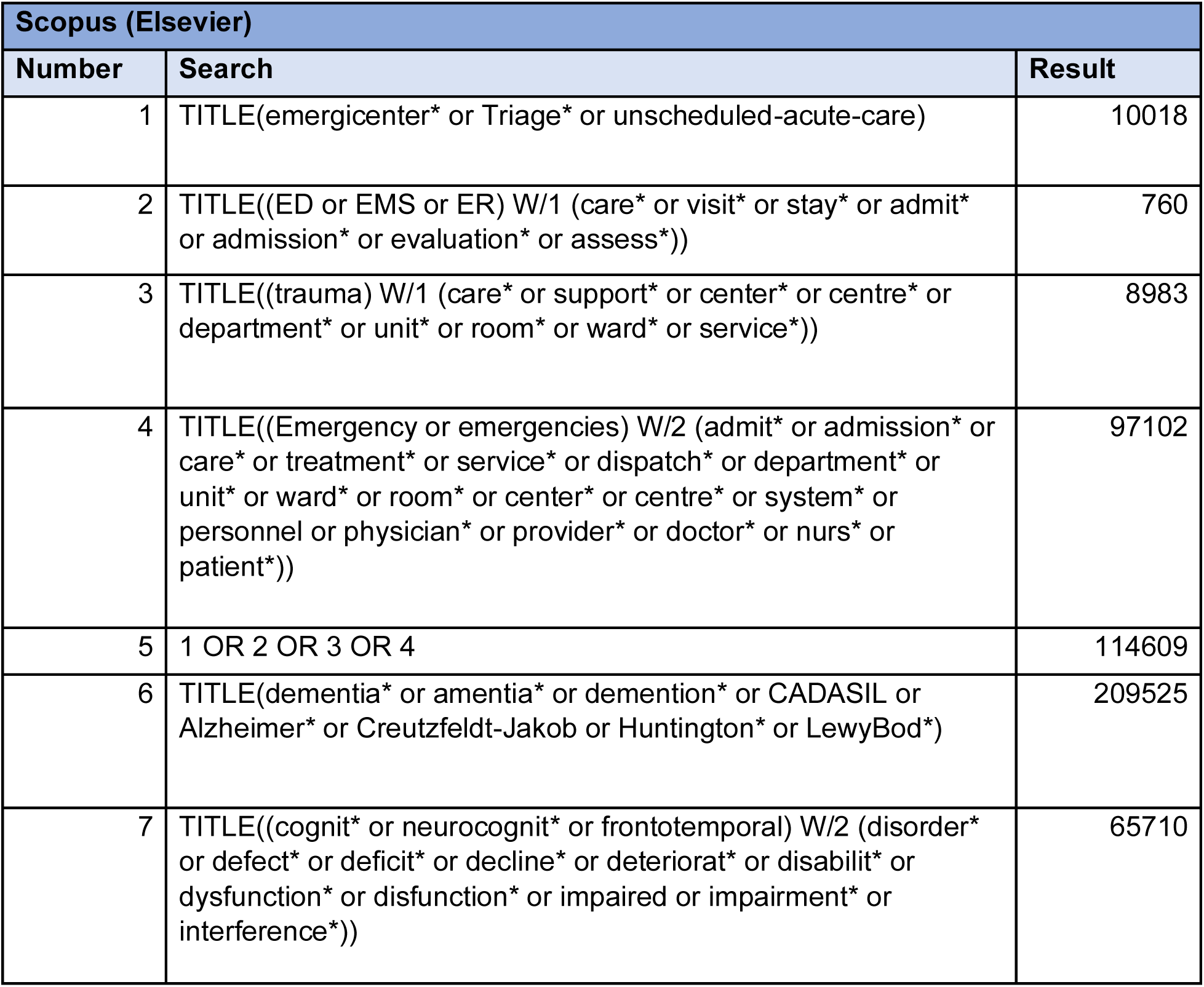
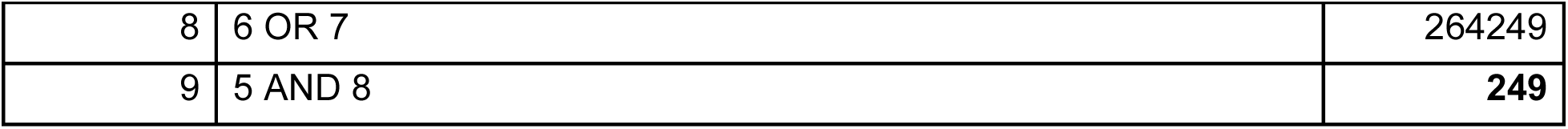
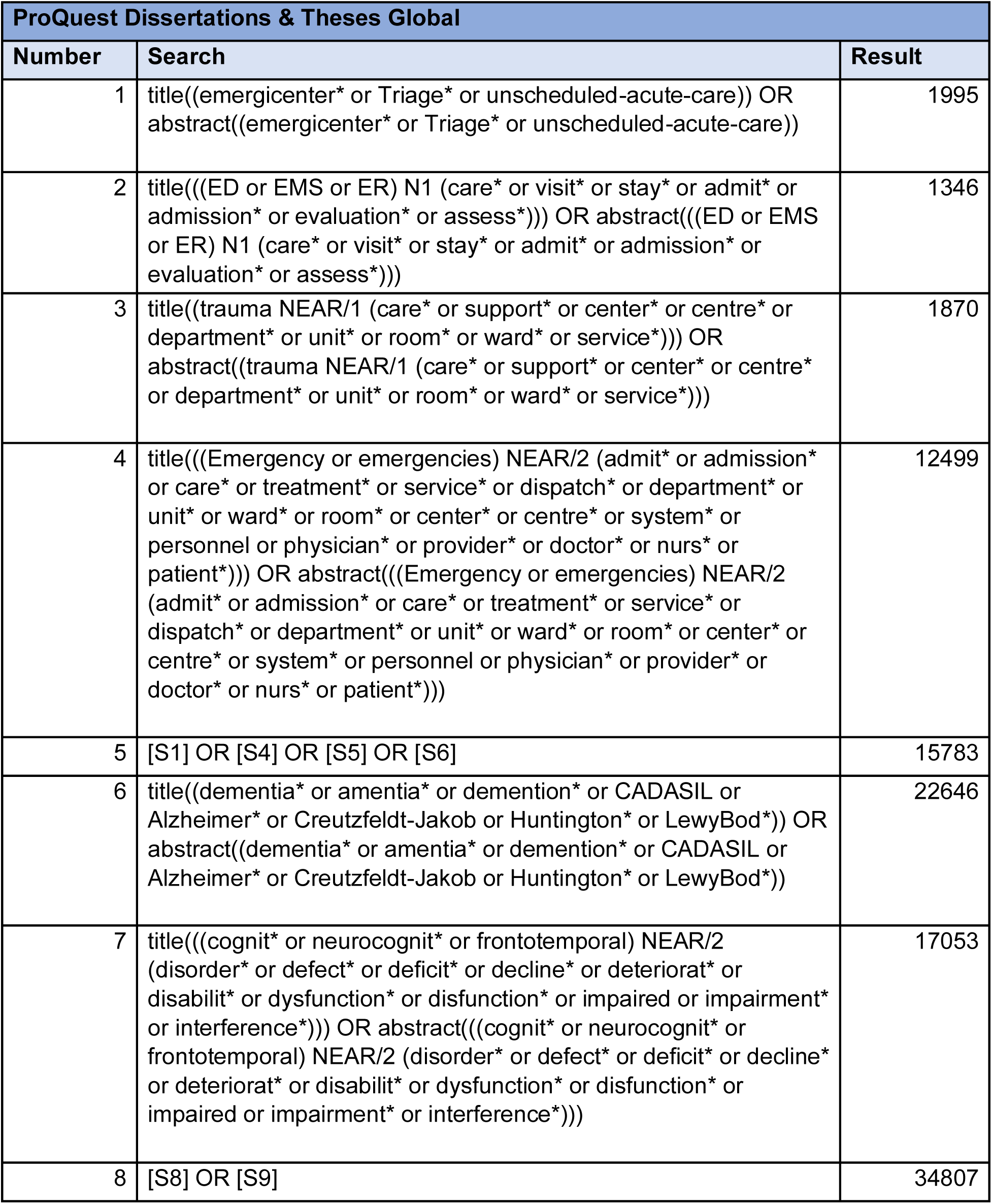
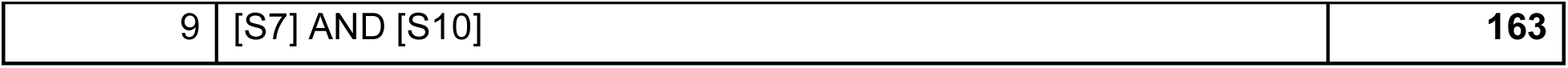

| Ovid MEDLINE(R) ALL <1946 to September 19, 2024> |  |  |
| --- | --- | --- |
| Number | Search | Results |
| 1 | exp Emergency Medical Services/ | 176906 |
| 2 | Emergency Medicine/ | 15651 |
| 3 | (emergicenter* or Triage* or unscheduled-acute-care).ti,ab. | 28213 |
| 4 | ((ED or EMS or ER) adj1 (care* or visit* or stay* or admit* or admission* or evaluation* or assess*)).ti,ab. | 18296 |
| 5 | (trauma adj1 (care* or support* or center* or centre* or department* or unit* or room* or ward* or service*)).ti,ab. | 30447 |
| 6 | ((Emergency or emergencies) adj2 (admit* or admission* or care* or treatment* or service* or dispatch* or department* or unit* or ward* or room* or center* or centre* or system* or personnel or physician* or provider* or doctor* or nurs* or patient*)):ti,ab. | 225629 |
| 7 | 1 or 2 or 3 or 4 or 5 or 6 | 347065 |
| 8 | exp Dementia/ | 219093 |
| 9 | (dementia* or amentia* or demention* or CADASIL or Alzheimer* or Creutzfeldt-Jakob or Huntington* or LewyBod*):ti,ab. | 317078 |
| 10 | ((cognit* or neurocognit* or frontotemporal) adj2 (disorder* or defect* or deficit* or decline* or deteriorat* or disabilit* or dysfunction* or disfunction* or impaired or impairment* or interference*)):ti,ab. | 193299 |
| 11 | 8 or 9 or 10 | 467038 |
| 12 | 7 and 11 | **2736** |

| **Cochrane Library (Wiley)** |  |  |
| --- | --- | --- |
| **Number** | **Search** | **Results** |
| #1 | MeSH descriptor: [Emergency Medical Services] explode all trees | 6186 |
| #2 | MeSH descriptor: [Emergency Medicine] explode all trees | 382 |
| #3 | ((emergicenter* or Triage* or unscheduled-acute-care)):ti,ab,kw | 2374 |
| #4 | ((ED or EMS or ER) NEAR/1 (care* or visit* or stay* or admit* or admission* or evaluation* or assess*)):ti,ab,kw | 2643 |
| #5 | ((trauma) NEAR/1 (care* or support* or center* or centre* or department* or unit* or room* or ward* or service*)):ti,ab,kw | 1840 |
| #6 | ((Emergency or emergencies) NEAR/2 (admit* or admission* or care* or treatment* or service* or dispatch* or department* or unit* or ward* or room* or center* or centre* or system* or personnel or physician* or provider* or doctor* or nurs* or patient*)):ti,ab,kw | 25519 |
| #7 | #1 OR #2 OR #3 OR #4 OR #5 OR #6 | 28872 |
| #8 | MeSH descriptor: [Dementia] explode all trees | 9590 |
| #9 | ((dementia* or amentia* or demention* or CADASIL or Alzheimer* or Creutzfeldt-Jakob or Huntington* or LewyBod*)):ti,ab,kw | 27259 |
| #10 | ((cognit* or neurocognit* or frontotemporal) NEAR/2 (disorder* or defect* or deficit* or decline* or deteriorat* or disabilit* or dysfunction* or disfunction* or impaired or impairment* or interference*)):ti,ab,kw | 30380 |
| #11 | #8 OR #9 OR #10 | 48960 |
| #12 | #7 AND #11 | **546** |

| Scopus (Elsevier) |  |  |
| --- | --- | --- |
| Number | Search | Result |
| 1 | TITLE(emergicenter* or Triage* or unscheduled-acute-care) | 10018 |
| 2 | TITLE((ED or EMS or ER) W/1 (care* or visit* or stay* or admit* or admission* or evaluation* or assess*)) | 760 |
| 3 | TITLE((trauma) W/1 (care* or support* or center* or centre* or department* or unit* or room* or ward* or service*)) | 8983 |
| 4 | TITLE((Emergency or emergencies) W/2 (admit* or admission* or care* or treatment* or service* or dispatch* or department* or unit* or ward* or room* or center* or centre* or system* or personnel or physician* or provider* or doctor* or nurs* or patient*)) | 97102 |
| 5 | 1 OR 2 OR 3 OR 4 | 114609 |
| 6 | TITLE(dementia* or amentia* or demention* or CADASIL or Alzheimer* or Creutzfeldt-Jakob or Huntington* or LewyBod*) | 209525 |
| 7 | TITLE((cognit* or neurocognit* or frontotemporal) W/2 (disorder* or defect* or deficit* or decline* or deteriorat* or disabilit* or dysfunction* or disfunction* or impaired or impairment* or interference*)) | 65710 |
| 8 | 6 OR 7 | 264249 |
| 9 | 5 AND 8 | **249** |

| ProQuest Dissertations & Theses Global |  |  |
| --- | --- | --- |
| Number | Search | Result |
| 1 | title((emergicenter* or Triage* or unscheduled-acute-care)) OR abstract((emergicenter* or Triage* or unscheduled-acute-care)) | 1995 |
| 2 | title(((ED or EMS or ER) N1 (care* or visit* or stay* or admit* or admission* or evaluation* or assess*))) OR abstract(((ED or EMS or ER) N1 (care* or visit* or stay* or admit* or admission* or evaluation* or assess*))) | 1346 |
| 3 | title((trauma NEAR/1 (care* or support* or center* or centre* or department* or unit* or room* or ward* or service*))) OR abstract((trauma NEAR/1 (care* or support* or center* or centre* or department* or unit* or room* or ward* or service*))) | 1870 |
| 4 | title(((Emergency or emergencies) NEAR/2 (admit* or admission* or care* or treatment* or service* or dispatch* or department* or unit* or ward* or room* or center* or centre* or system* or personnel or physician* or provider* or doctor* or nurs* or patient*))) OR abstract(((Emergency or emergencies) NEAR/2 (admit* or admission* or care* or treatment* or service* or dispatch* or department* or unit* or ward* or room* or center* or centre* or system* or personnel or physician* or provider* or doctor* or nurs* or patient*))) | 12499 |
| 5 | [S1] OR [S4] OR [S5] OR [S6] | 15783 |
| 6 | title((dementia* or amentia* or demention* or CADASIL or Alzheimer* or Creutzfeldt-Jakob or Huntington* or LewyBod*)) OR abstract((dementia* or amentia* or demention* or CADASIL or Alzheimer* or Creutzfeldt-Jakob or Huntington* or LewyBod*)) | 22646 |
| 7 | title(((cognit* or neurocognit* or frontotemporal) NEAR/2 (disorder* or defect* or deficit* or decline* or deteriorat* or disabilit* or dysfunction* or disfunction* or impaired or impairment* or interference*))) OR abstract(((cognit* or neurocognit* or frontotemporal) NEAR/2 (disorder* or defect* or deficit* or decline* or deteriorat* or disabilit* or dysfunction* or disfunction* or impaired or impairment* or interference*))) | 17053 |
| 8 | [S8] OR [S9] | 34807 |
| 9 | [S7] AND [S10] | 163 |

## Appendix 2. PRISMA checklist for manuscript

Appendix 2. PRISMA checklist for manuscript.
| Section and Topic | Item # | Checklist item | Location where item is reported |
| --- | --- | --- | --- |
| **TITLE** |  |  |  |
| Title | 1 | Identify the report as a systematic review. | Yes |
| **ABSTRACT** |  |  |  |
| Abstract | 2 | See the PRISMA 2020 for Abstracts checklist. | Filled out |
| **INTRODUCTION** |  |  |  |
| Rationale | 3 | Describe the rationale for the review in the context of existing knowledge. | Pg 3 |
| Objectives | 4 | Provide an explicit statement of the objective(s) or question(s) the review addresses. | Pg 3 |
| **METHODS** |  |  |  |
| Eligibility criteria | 5 | Specify the inclusion and exclusion criteria for the review and how studies were grouped for the syntheses. |  |
| Information sources | 6 | Specify all databases, registers, websites, organisations, reference lists and other sources searched or consulted to identify studies. Specify the date when each source was last searched or consulted. | Pg 4, edits made |
| Search strategy | 7 | Present the full search strategies for all databases, registers and websites, including any filters and limits used. | Appendix 1 |
| Selection process | 8 | Specify the methods used to decide whether a study met the inclusion criteria of the review, including how many reviewers screened each record and each report retrieved, whether they worked independently, and if applicable, details of automation tools used in the process. | Pg 4 |
| Data collection process | 9 | Specify the methods used to collect data from reports, including how many reviewers collected data from each report, whether they worked independently, any processes for obtaining or confirming data from study investigators, and if applicable, details of automation tools used in the process. | Pg 5 |
| Data items | 10a | List and define all outcomes for which data were sought. Specify whether all results that were compatible with each outcome domain in each study were sought (e.g. for all measures, time points, analyses), and if not, the methods used to decide which results to collect. | Pg 5 |
|  | 10<br>b | List and define all other variables for which data were sought (e.g. participant and intervention characteristics, funding sources). Describe any assumptions made about any missing or unclear information. | Pg 5 |
| Study risk of bias assessment | 11 | Specify the methods used to assess risk of bias in the included studies, including details of the tool(s) used, how many reviewers assessed each study and whether they worked independently, and if applicable, details of automation tools used in the process. | Pg 5 |
| Effect measures | 12 | Specify for each outcome the effect measure(s) (e.g. risk ratio, mean difference) used in the synthesis or presentation of results. | Pg 5 |
| Synthesis methods | 13<br>a | Describe the processes used to decide which studies were eligible for each synthesis (e.g. tabulating the study intervention characteristics and comparing against the planned groups for each synthesis (item #5)). | Pg 5 |
|  | 13<br>b | Describe any methods required to prepare the data for presentation or synthesis, such as handling of missing summary statistics, or data conversions. | Pg 5 |
|  | 13<br>c | Describe any methods used to tabulate or visually display results of individual studies and syntheses. | Tables /figures |
|  | 13<br>d | Describe any methods used to synthesize results and provide a rationale for the choice(s). If meta-analysis was performed, describe the model(s), method(s) to identify the presence and extent of statistical heterogeneity, and software package(s) used. | n/a |
|  | 13<br>e | Describe any methods used to explore possible causes of heterogeneity among study results (e.g. subgroup analysis, meta-regression). | n/a |
|  | 13<br>f | Describe any sensitivity analyses conducted to assess robustness of the synthesized results. | n/a |
| Reporting bias assessment | 14 | Describe any methods used to assess risk of bias due to missing results in a synthesis (arising from reporting biases). | Figure 2, Figure 3 |
| Certainty assessment | 15 | Describe any methods used to assess certainty (or confidence) in the body of evidence for an outcome. | n/a |
| **RESULTS** |  |  |  |
| Study selection | 16 a | Describe the results of the search and selection process, from the number of records identified in the search to the number of studies included in the review, ideally using a flow diagram. | Pg 6, Figure 1 |
|  | 16 b | Cite studies that might appear to meet the inclusion criteria, but which were excluded, and explain why they were excluded. | Fig 1 |
| Study characteristics | 17 | Cite each included study and present its characteristics. | Pg 6-7 |
| Risk of bias in studies | 18 | Present assessments of risk of bias for each included study. | Pg 6-7, Fig 2&3 |
| Results of individual studies | 19 | For all outcomes, present, for each study: (a) summary statistics for each group (where appropriate) and (b) an effect estimate and its precision (e.g. confidence/credible interval), ideally using structured tables or plots. | n/a |
| Results of syntheses | 20 a | For each synthesis, briefly summarise the characteristics and risk of bias among contributing studies. | Pg 8 |
|  | 20 b | Present results of all statistical syntheses conducted. If meta-analysis was done, present for each the summary estimate and its precision (e.g. confidence/credible interval) and measures of statistical heterogeneity. If comparing groups, describe the direction of the effect. | n/a |
|  | 20 c | Present results of all investigations of possible causes of heterogeneity among study results. | n/a |
|  | 20 d | Present results of all sensitivity analyses conducted to assess the robustness of the synthesized results. | n/a |
| Reporting biases | 21 | Present assessments of risk of bias due to missing results (arising from reporting biases) for each synthesis assessed. | n/a |
| Certainty of evidence | 22 | Present assessments of certainty (or confidence) in the body of evidence for each outcome assessed. | n/a |
| **DISCUSSION** |  |  |  |
| Discussion | 23 a | Provide a general interpretation of the results in the context of other evidence. | Pg 8 & 9 |
|  | 23 b | Discuss any limitations of the evidence included in the review. | Pg 8 & 9 |
|  | 23<br>c | Discuss any limitations of the review processes used. | Pg 9 |
|  | 23<br>d | Discuss implications of the results for practice, policy, and future research. | Pg 10 |
| **OTHER INFORMATION** |  |  |  |
| Registrati<br>on and<br>protocol | 24<br>a | Provide registration information for the review, including register name and registration number, or state that the review was not registered. | Yes |
|  | 24<br>b | Indicate where the review protocol can be accessed, or state that a protocol was not prepared. | Yes |
|  | 24<br>c | Describe and explain any amendments to information provided at registration or in the protocol. | n/a |
| Support | 25 | Describe sources of financial or non-financial support for the review, and the role of the funders or sponsors in the review. | Not<br>stated |
| Competin<br>g<br>interests | 26 | Declare any competing interests of review authors. | Pg 3 |
| Availabilit<br>y of data,<br>code and<br>other<br>materials | 27 | Report which of the following are publicly available and where they can be found: template data collection forms; data extracted from included studies; data used for all analyses; analytic code; any other materials used in the review. | Includ<br>ed |

*From:* Page MJ, McKenzie JE, Bossuyt PM, Boutron I, Hoffmann TC, Mulrow CD, et al. The PRISMA 2020 statement: an updated guideline for reporting systematic reviews. *BMJ* 2021;372:n71. doi: 10.1136/bmj.n71. This work is licensed under CC BY 4.0. To view a copy of this license, visit https://creativecommons.org/licenses/by/4.0/

## Appendix 3. PRISMA checklist for abstract

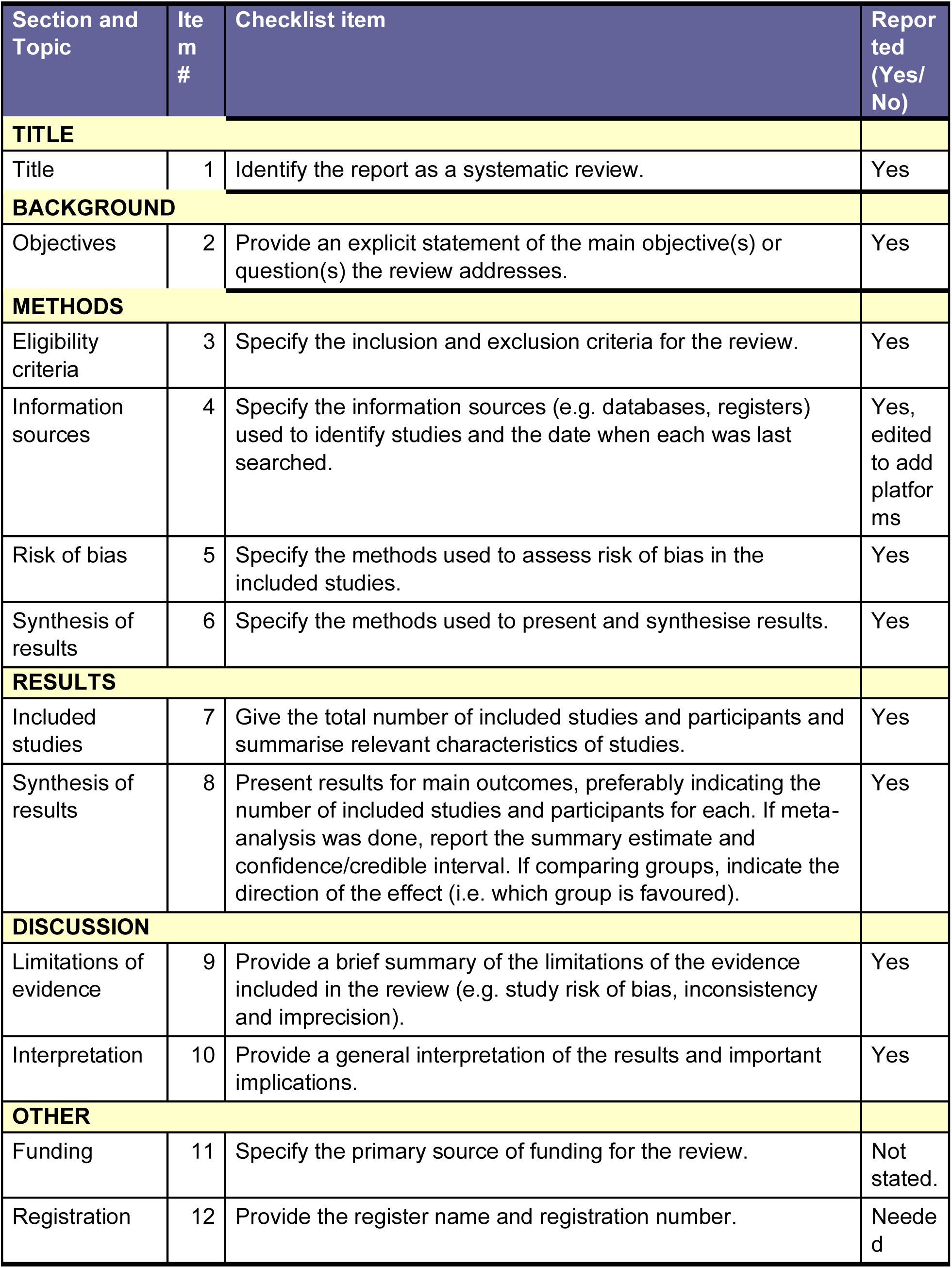
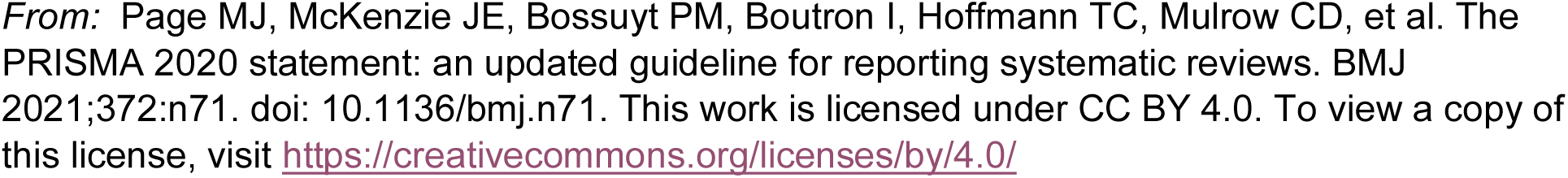

## Appendix 4. New-Castle Ottawa Scale

**NEWCASTLE - OTTAWA QUALITY ASSESSMENT SCALE**

**COHORT STUDIES**^58^

<u>Note</u>: A study can be awarded a maximum of one star for each numbered item within the Selection and Outcome categories. A maximum of three stars can be given for Comparability Max score of 9. 0-3 high risk, 4-7 moderate, 8-9 high risk of bias

### Selection

1) <u>Representativeness of the exposed cohort (0 to 1)</u>

a) truly representative of the average ED patients with dementia in the real world*
b) somewhat representative of the average ED patients with dementia in the real world*
c) selected group of users eg older adults, volunteers
d) no description of the derivation of the cohort
Dementia defined by past history of dementia, Alzheimer’s disease and related dementia (ADRD), or proxy history of dementia/ADRD, risk stratification tool, or dementia screening/assessment tools, and this is not a new diagnosis made by ED clinicians
2) <u>Selection of the non-exposed cohort (0 to 1)</u>

a) drawn from the same community (ED patients with dementia) as the exposed cohort*
b) drawn from a different source (For example, ED patients without dementia)
c) no description of the derivation of the non-exposed cohort
3) <u>Ascertainment of exposure (0 to 1 for either a or b)</u>

a) structured intervention*
b) secure record (eg surgical records verifying the receipt of intervention, not counting generic EMR documents)*
c) self report
d) no description
4) <u>Demonstration that outcome of interest was not present at start of study, in other words, there is a change of status measured (or not measured) at the endpoint (0 to 1)</u>

a) yes*
b) no

### Comparability

1) <u>Comparability of cohorts on the basis of the design or analysis (0 to 3)</u>
  a) study controls for stage of dementia/frailty
  b) study controls for any additional factor of triage acuity (example, Emergency Severity Index or ESI)
  c) study control for additional factor of place of living

### Outcome

1) <u>Assessment of outcome</u> (0 to 1 for a or b)
  a) independent blind assessment*
  b) record linkage*
  c) self-report
  d) no description
2) <u>Was follow-up long enough for outcomes to occur (0 to 1)</u>

a) yes (select an adequate follow up period for outcome of interest-3 months)
b) no
3) <u>Adequacy of follow up of cohorts (0 to 1 for either a or b)</u>

a) complete follow up - all subjects accounted for*
b) subjects lost to follow up unlikely to introduce bias - small number lost - > 20 % (select an adequate %) follow up, or description provided of those lost)*
c) follow up rate < 20% (select an adequate %) and no description of those lost
d) no statement

## Appendix 5. Cochrane risk of bias (2.0) tool (for parallel arm)

### Risk of bias assessment ^59^

Responses <u>underlined in green</u> are potential markers for low risk of bias, and responses in red are potential markers for a risk of bias. Where questions relate only to sign posts to other questions, no formatting is used.

**Domain 1:** Risk of bias arising from the randomization process.

| Signalling questions | Comments | Response options |
| --- | --- | --- |
| 1.1 Was the allocation sequence random? |  | <u>Y / PY</u> / **PN / N** / NI |
| 1.2 Was the allocation sequence concealed until participants were enrolled and assigned to interventions? |  | <u>Y / PY</u> / **PN / N** / NI |
| 1.3 Did baseline differences between intervention groups suggest a problem with the randomization process? |  | **Y / PY** / <u>PN / N</u> / NI |
| Risk-of-bias judgement | • | Low / High / Some concerns |
| Optional: What is the predicted direction of bias arising from the randomization process? |  | NA / Favours experimental / Favours comparator / Towards null / Away from null / Unpredictable |

**Domain 2:**
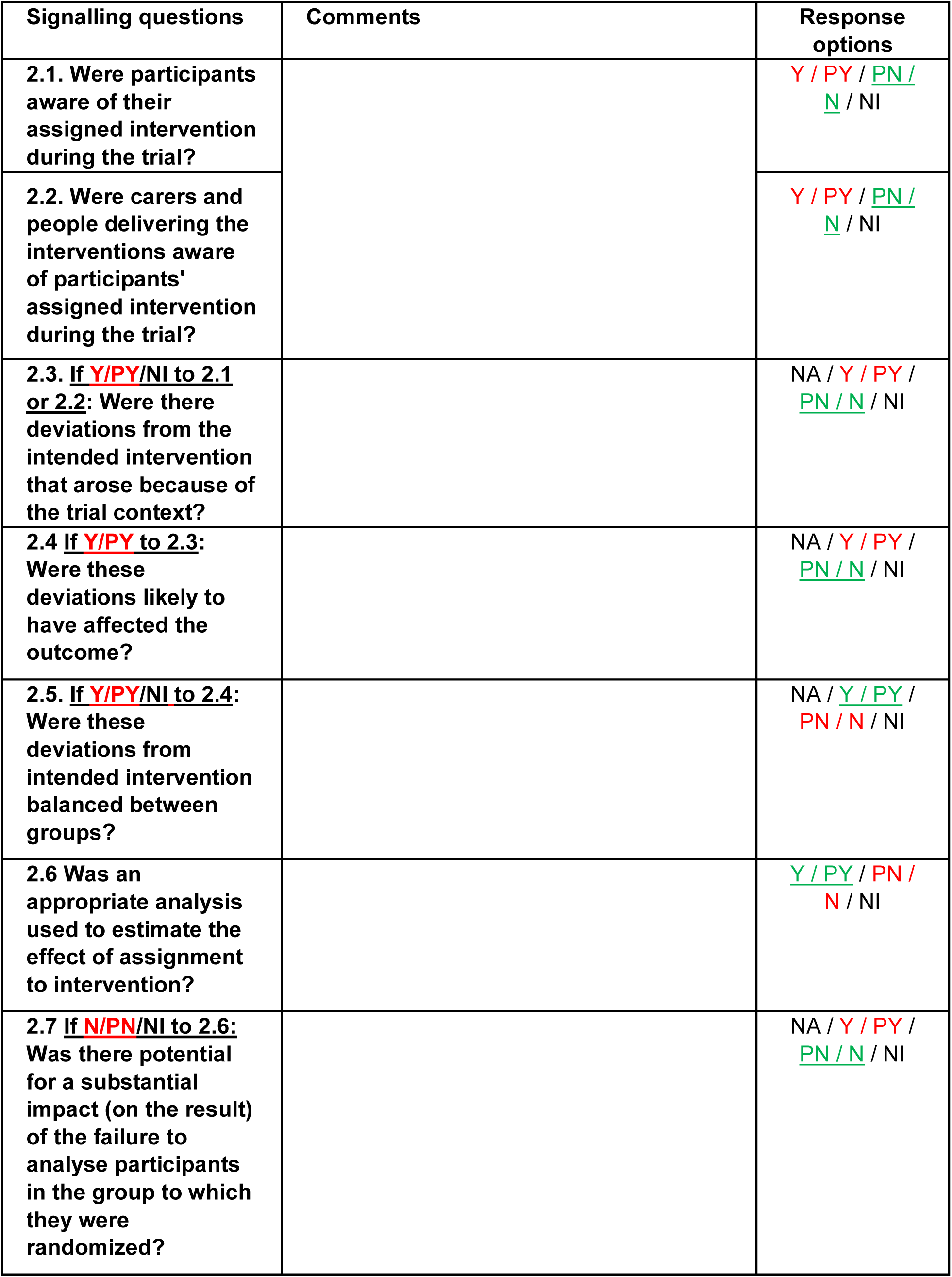
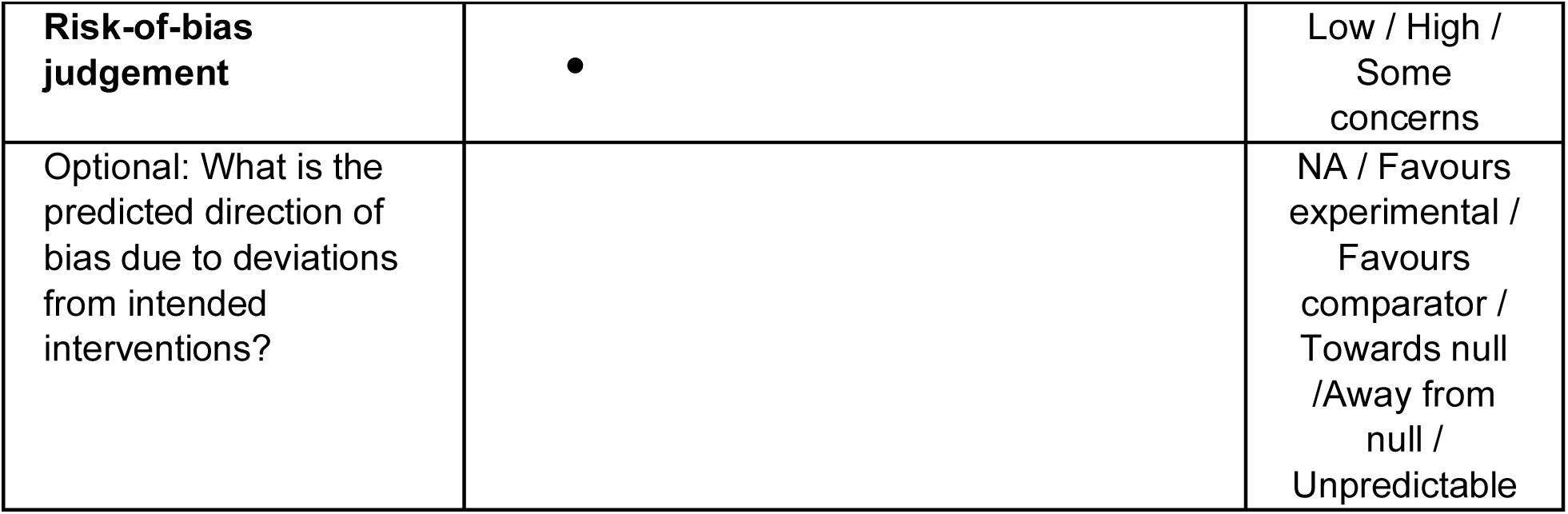
Risk of bias due to deviations from the intended interventions (*effect of assignment to intervention*)

| Signalling questions | Comments | Response options |
| --- | --- | --- |
| 2.1. Were participants aware of their assigned intervention during the trial? |  | Y / PY / <u>PN</u> / <u>N</u> / NI |
| 2.2. Were carers and people delivering the interventions aware of participants' assigned intervention during the trial? |  | Y / PY / <u>PN</u> / <u>N</u> / NI |
| 2.3. If <u>Y/PY/NI</u> to 2.1 or 2.2: Were there deviations from the intended intervention that arose because of the trial context? |  | NA / Y / PY / <u>PN</u> / <u>N</u> / NI |
| 2.4 If <u>Y/PY</u> to 2.3: Were these deviations likely to have affected the outcome? |  | NA / Y / PY / <u>PN</u> / <u>N</u> / NI |
| 2.5. If <u>Y/PY/NI</u> to 2.4: Were these deviations from intended intervention balanced between groups? |  | NA / <u>Y</u> / PY / <u>PN</u> / <u>N</u> / NI |
| 2.6 Was an appropriate analysis used to estimate the effect of assignment to intervention? |  | <u>Y</u> / PY / <u>PN</u> / <u>N</u> / NI |
| 2.7 If <u>N/PN/NI</u> to 2.6: Was there potential for a substantial impact (on the result) of the failure to analyse participants in the group to which they were randomized? |  | NA / Y / PY / <u>PN</u> / <u>N</u> / NI |
| **Risk-of-bias judgement** | • | Low / High /<br>Some concerns |
| Optional: What is the predicted direction of bias due to deviations from intended interventions? |  | NA / Favours experimental /<br>Favours comparator /<br>Towards null<br>/Away from null /<br>Unpredictable |

**Domain 2:**
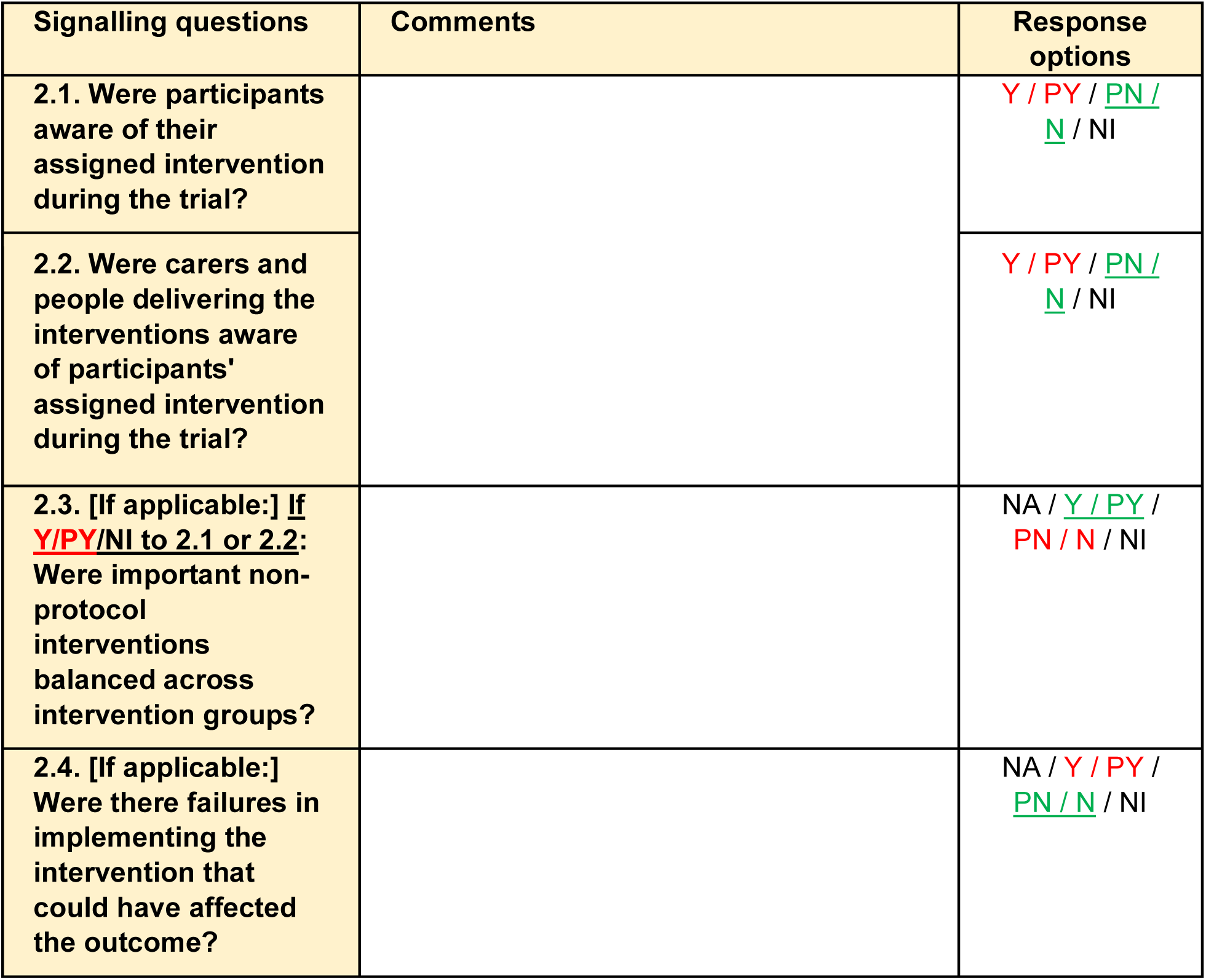
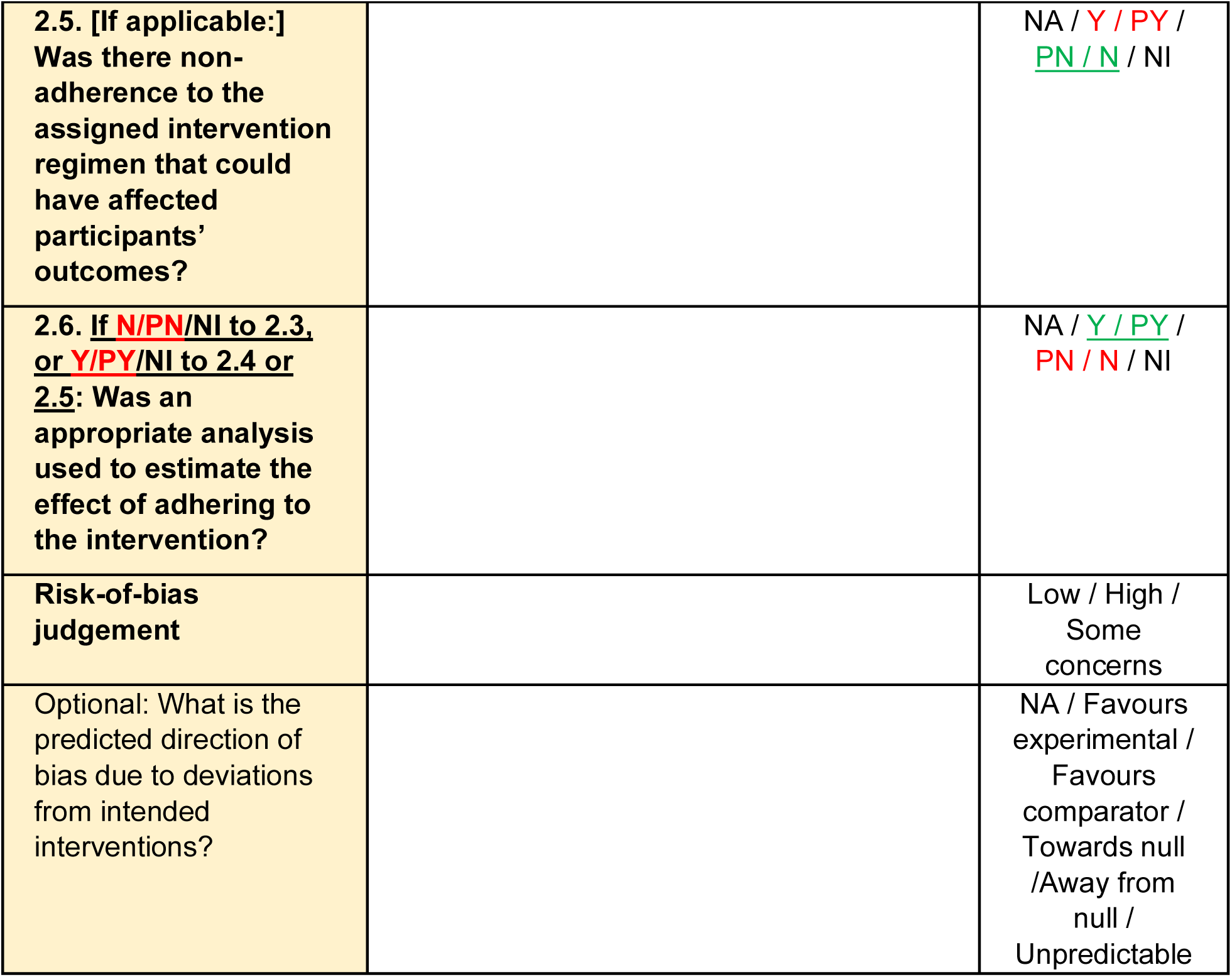
Risk of bias due to deviations from the intended interventions (*effect of adhering to intervention*)

**Domain 3:** Missing outcome data.

| **Signalling questions** | **Comments** | **Response options** |
| --- | --- | --- |
| **3.1** Were data for this outcome available for all, or nearly all, participants randomized? |  | **Y** / **PY** / **PN** / **N** / NI |
| **3.2** If **N/PN/NI** to 3.1: Is there evidence that the result was not biased by missing outcome data? |  | NA / **Y** / **PY** / **PN** / **N** |
| **3.3 If <u>N/PN</u> to 3.2:**<br>Could missingness in the outcome depend on its true value? |  | NA / **Y** / **PY** / <u>PN / N</u> / NI |
| **3.4 If <u>Y/PY/NI</u> to 3.3:**<br>Is it likely that missingness in the outcome depended on its true value? |  | NA / **Y** / **PY** / <u>PN / N</u> / NI |
| **Risk-of-bias judgement** |  | Low / High / Some concerns |
| Optional: What is the predicted direction of bias due to missing outcome data? |  | NA / Favours experimental / Favours comparator / Towards null / Away from null / Unpredictable |

**Domain 4:** Risk of bias in measurement of the outcome.

| **Signalling questions** | **Comments** | **Response options** |
| --- | --- | --- |
| **4.1 Was the method of measuring the outcome inappropriate?** |  | **Y** / **PY** / <u>PN / N</u> / NI |
| **4.2 Could measurement or ascertainment of the outcome have differed between intervention groups?** |  | **Y** / **PY** / <u>PN / N</u> / NI |
| **4.3 If <u>N/PN/NI</u> to 4.1 and 4.2: Were outcome assessors aware of the intervention received** |  | NA / **Y** / **PY** / <u>PN / N</u> / NI |
| by study participants? |  |  |
| 4.4 If **Y/PY/NI** to 4.3:<br>Could assessment of the outcome have been influenced by knowledge of intervention received? |  | NA / **Y** / **PY** / **PN** / **N** / NI |
| 4.5 If **Y/PY/NI** to 4.4:<br>Is it likely that assessment of the outcome was influenced by knowledge of intervention received? |  | NA / **Y** / **PY** / **PN** / **N** / NI |
| Risk-of-bias judgement |  | Low / High / Some concerns |
| Optional: What is the predicted direction of bias in measurement of the outcome? |  | NA / Favours experimental / Favours comparator / Towards null / Away from null / Unpredictable |

**Domain 5:**
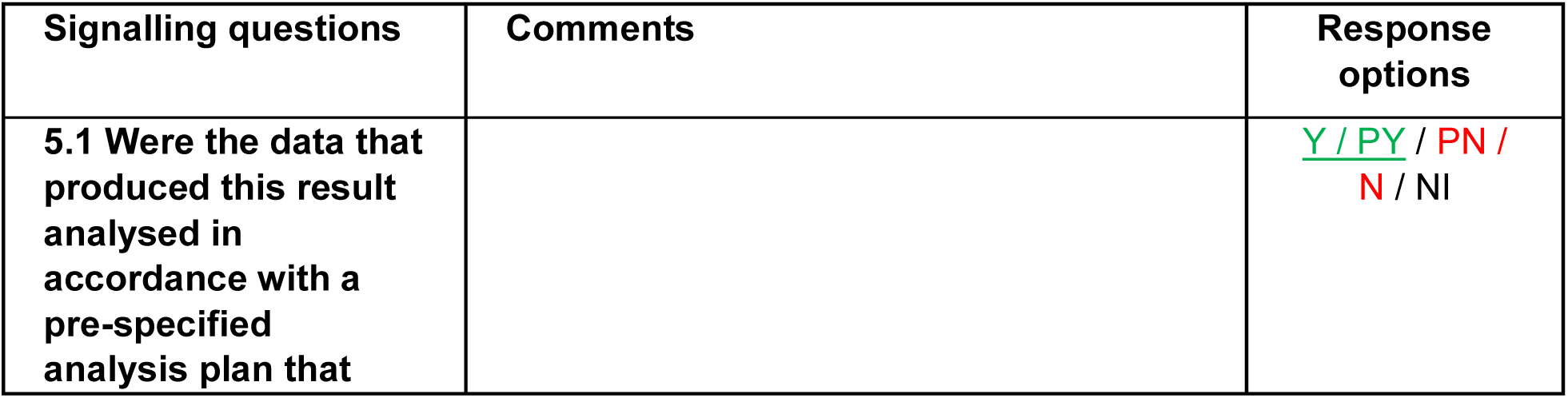
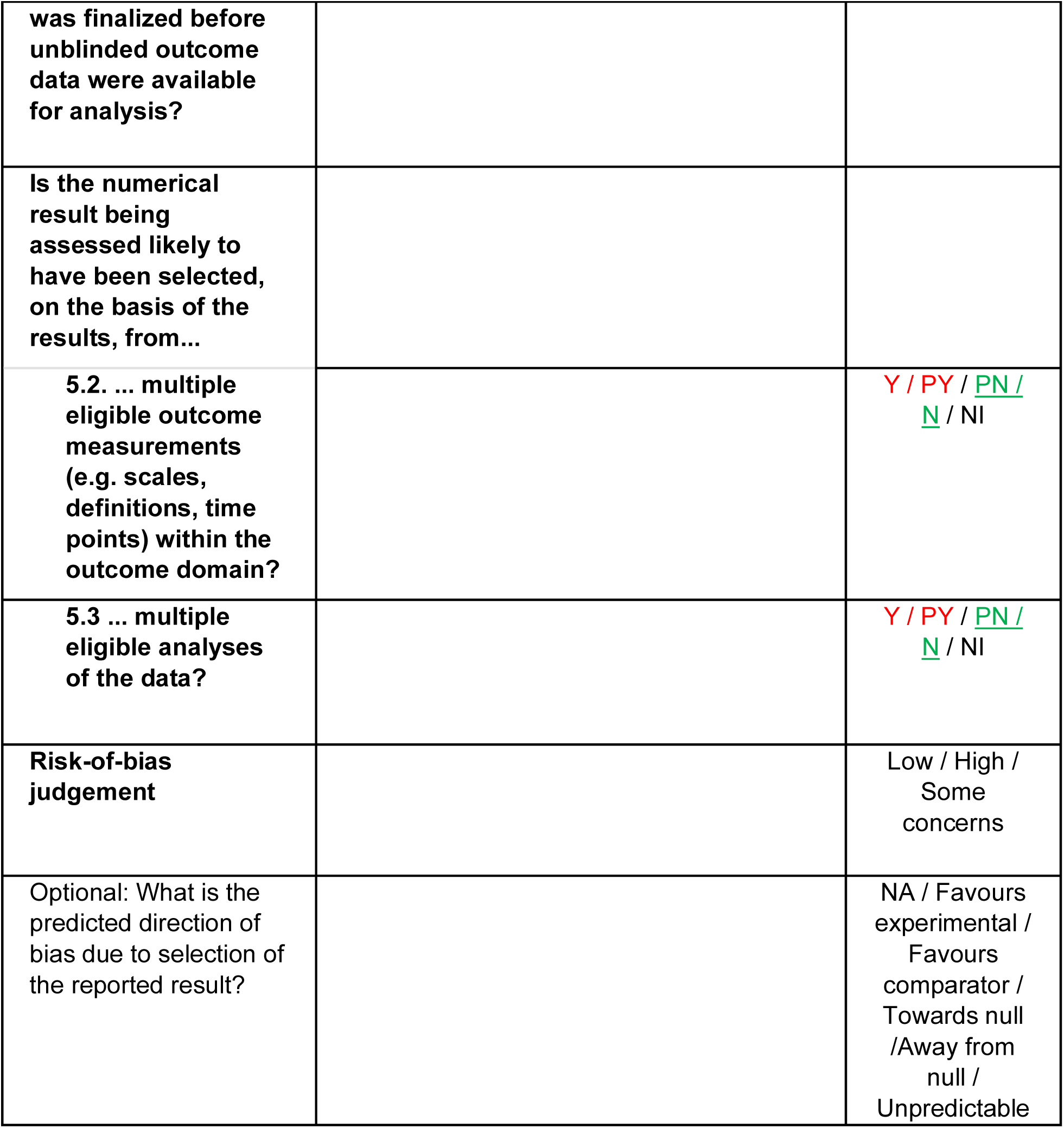
Risk of bias in selection of the reported result.

| Signalling questions | Comments | Response options |
| --- | --- | --- |
| 5.1 Were the data that produced this result analysed in accordance with a pre-specified analysis plan that |  | **Y** / **PY** / **PN** / **N** / NI |
| was finalized before unblinded outcome data were available for analysis? |  |  |
| Is the numerical result being assessed likely to have been selected, on the basis of the results, from... |  |  |
| 5.2. ... multiple eligible outcome measurements (e.g. scales, definitions, time points) within the outcome domain? |  | Y / PY / <u>PN</u> / <u>N</u> / NI |
| 5.3 ... multiple eligible analyses of the data? |  | Y / PY / <u>PN</u> / <u>N</u> / NI |
| Risk-of-bias judgement |  | Low / High / Some concerns |
| Optional: What is the predicted direction of bias due to selection of the reported result? |  | NA / Favours experimental / Favours comparator / Towards null / Away from null / Unpredictable |

### Overall risk of bias

|  |  |  |
| --- | --- | --- |
| **Risk-of-bias judgement** |  | Low / High /<br>Some concerns |
| Optional: What is the overall predicted direction of bias for this outcome? |  | NA / Favours experimental / Favours comparator / Towards null / Away from null / Unpredictable |

This work is licensed under a [Creative Commons Attribution-NonCommercial-NoDerivatives 4.0 International License](http://creativecommons.org/licenses/by-nc-nd/4.0/).

Geriatric ED Guidelines dementia writing group

Scott Dresden, MD
Associate Professor
Department of Emergency Medicine
Northwestern University Feinberg School of Medicine
Chicago, IL

Angel Li MD, MBA
Assistant Professor
Department of Emergency Medicine,
The Ohio State University

Alexander X. Lo, MD, PhD
Associate Professor
Department of Emergency Medicine
Northwestern University Feinberg School of Medicine
Chicago, IL

Cameron J. Gettel, MD MHS
Assistant Professor
Department of Emergency Medicine
Yale School of Medicine
New Haven, CT

R. Doreen Monks, RN (retired), MSN
Board Member
Voice of Alzheimer’s
Washington DC

